# Preclinical and Human Phase 1 Studies of Aerosolized Hydroxychloroquine: Implications for Antiviral COVID-19 Therapy

**DOI:** 10.1101/2023.06.22.23291702

**Authors:** Ohad S. Bentur, Richard Hutt, Donna Brassil, Ana C. Kriegegr, Per Bäckman, B. Lauren Charous, Homer Boushey, Igor Gonda, Barry S. Coller, Robert B. MacArthur

**Author notes:** Correspondence: Robert B. MacArthur, The Rockefeller University Hospital, 1230 York Avenue, New York, NY, 10065.

## Abstract

Based on early reports of the efficacy of hydroxychloroquine sulfate (HCQS) to inhibit SARS-CoV-2 viral replication in vitro, and since severe pulmonary involvement is the major cause of COVID-19 mortality, we assessed the safety and efficacy of aerosolized HCQS (aHCQS) therapy in animals and humans. In a Phase 1 study of aHCQS in healthy volunteers, doses up to 50 mg were well tolerated and estimated epithelial lining fluid concentrations immediately after inhalation (>2,000 μM) exceeded the in vitro concentrations needed for suppression of viral replication (≥119 μM). A study in rats comparing HCQS solution administered orally (13.3 mg/kg) and by intratracheal installation (IT 0.18 mg/kg, <5% of oral dose) demonstrated that at 2 minutes, IT administration was associated with 5X higher mean hydroxychloroquine (HCQ) concentrations in the lung (IT: 49.5 ± 6.5 µg HCQ/g tissue, oral: 9.9 ± 3.4; p<0.01). A subsequent study of IT and intranasal HCQS in the Syrian hamster model of SARS-CoV-2 infection, however, failed to show clinical benefit. We conclude that aHCQS alone is unlikely to be effective for COVID-19, but based on our aHCQS pharmacokinetics and current viral entry data, adding oral HCQS to aHCQS, along with a transmembrane protease inhibitor, may improve efficacy.

## Introduction

The first 4 cases of what would later be called SARS-CoV-2 infection, or COVID-19, were reported in Wuhan, China on December 31, 2019 (1, 2), and by January 31, 2020 the World Health Organization (WHO) declared the disease a Global Health Emergency (3) and the United States (U.S.) declared the disease a Public Health Emergency (4). On March 28, 2020, the U.S. Food and Drug Administration (FDA) issued an Emergency Use Authorization (EUA) for hydroxychloroquine sulfate (HCQS), noting in vitro and anecdotal in vivo data, concluding that “Based on the totality of scientific evidence available to FDA, it is reasonable to believe that…HCQS may be effective in treating COVID-19” (5). However, on June 15, 2020 the FDA revoked the EUA after determining that HCQS is “unlikely to be effective in treating COVID-19” (6).

HCQS is a commonly used anti-malarial and anti-rheumatic agent that may have a notably prolonged onset of action due to its large volume of distribution (7). One week before the FDA issued the EUA authorizing the use of HCQS, and based on reports of the ability of HCQS to inhibit SARS-CoV and SARS-CoV-2 replication in vitro in several different cell lines (8–10) that were later supported by additional studies (Supplementary Table 1), as well as an early study suggesting its clinical efficacy (11), one of the authors raised the idea of using aerosolized HCQS (aHCQS) as a means to rapidly achieve high concentrations of hydroxychloroquine (HCQ) in the airways and lungs, the primary targets of SARS-CoV-2, while minimizing the risk of cardiac toxicity, the major safety concern with orally administered HCQS (oHCQS) (12), especially in light of initial reports of the cardiac effects of COVID-19 (13). Several of the authors previously participated in conducting preclinical studies of aHCQS, a Phase 1 study of aHCQS in healthy volunteers, and then a Phase 2 study of aHCQS as a treatment for asthma, in which the drug was well tolerated at doses up to 20 mg given daily for 21 days, although it failed to meet the study primary objective (e.g. lead to a significant treatment related improvement in forced expiratory volume in 1 second (FEV_1),_with baseline correction) (14–17). Based on these considerations, the authors proceeded to develop a formulation for aHCQS, selected a nebulizer that would selectively target the airways as well as the lungs, and received regulatory agreement from the FDA to proceed to a Phase 1 study in healthy participants to assess the safety, tolerability and pharmacokinetics (PK) of aHCQS. Enrollment in that study started in June and ended in August 2020.

Published data on the PK of oHCQS were limited with regard to lung versus systemic distribution, although early work indicated that HCQ is retained in well-perfused organs including the lung (18, 19). Therefore, modeling of an effective clinical dose based upon published in vitro data presented challenges. The very large volume of distribution, in one report described as 2,851 ± 2,147 liters (20), and the very long terminal half-life of oHCQS, on the order of months (12), added to the complexity of modeling different doses. Further adding to the complexity is the wide range of values reported to achieve 50% inhibition of viral replication in cell cultures, some of which reflect the cell type used in the assays (Supplementary Table 1). An additional complexity relates to both the uncertainty of the mechanism by which HCQ inhibits viral replication, and that the virus can bypass HCQ antiviral mechanisms, depending upon the cell entry pathway followed (Figure 1).

**Figure 1.**
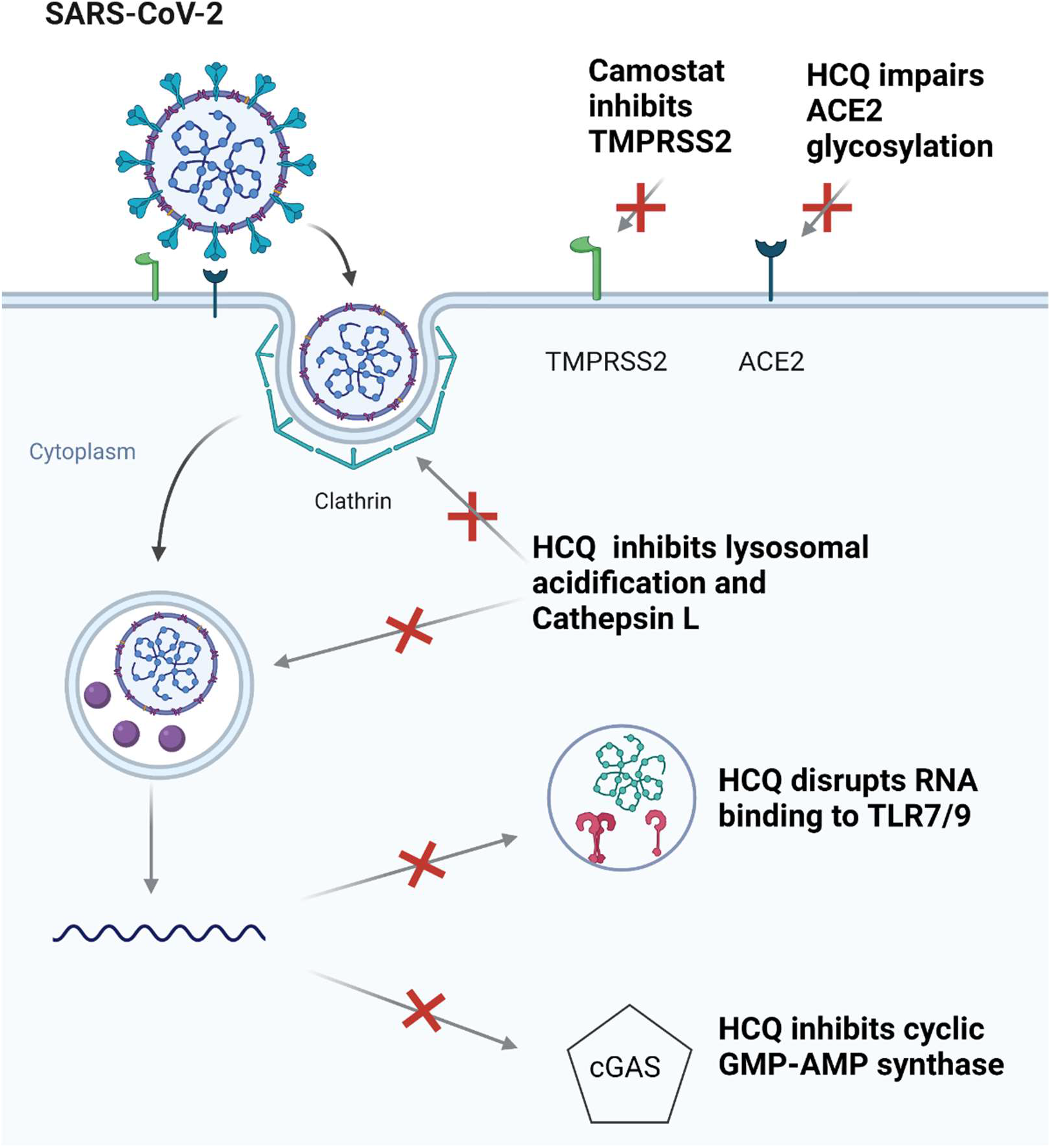
Proposed intracellular and extracellular sites of HCQ and TMPRSS2 inhibitor antiviral activity. The site of action determines where effective free drug concentrations must be achieved. Intracellular HCQ causes lysosomal alkalinization, which disrupts viral entry, unfolding, and reproduction. HCQ intracellular immunomodulatory effects include inhibition of Toll-like receptor activation and signaling via inhibition of cGAS. Extracellularly, TMPRSS2 inhibitors (camostat mesylate, nafamostat mesylate, others), block TMPRSS2 mediated viral entry. HCQ may also interfere with N-terminal glycosylation of ACE2 (54, 57, 59).

One hypothesis focuses upon the recognized intracellular pharmacological actions of HCQ, in which case the intracellular concentration is most important. Intracellular HCQ causes both lysosomal alkalinization and immunomodulatory effects, with the former disrupting intracellular viral unfolding, entry, and reproduction, and the latter mediated by inhibition of Toll-like receptor activation and signaling through cGAS mechanisms. Another hypothesis focuses on effects at the surface of the cell, in particular N-terminal glycosylation of angiotensin converting enzyme 2 (ACE2), in which case the extracellular free drug concentration in plasma may be most important (21–24).

HCQS and its analogue, chloroquine phosphate, distribute selectively in tissues, including blood cells, which accounts for the higher whole blood concentrations than plasma concentrations (7, 18, 19, 25–28). To gain a better understanding of the PK and tissue distribution of HCQ, we conducted a study comparing oral (PO), intravenous (IV), and intratracheal (IT) administration of HCQS solution in rats.

During the development of the aHCQS Phase 1 study, data began to accumulate challenging the efficacy of oHCQS in COVID-19 (29–33), but leaving open the possibility that aHCQS may still be effective by achieving higher lung levels faster than can be achieved with oHCQS at doses that do not produce unacceptable toxicity. In parallel, the Golden Syrian hamster model of SARS-CoV-2 infection emerged as a robust preclinical model and indicator of the efficacy of vaccines and medications (34–37). We therefore proceeded to conduct a study of IT and intranasal (IN) HCQS solution in this model to assess the potential efficacy of aHCQS.

## Results

### Preclinical PK and tissue distribution study of HCQ after single PO, IV, and IT administration of HCQS solution in male Sprague Dawley rats

Twenty animals were studied as indicated in Supplementary Table 2. Different total doses for each route of administration were selected based on the conventions of allometric scaling of inhaled drugs (see Supplementary Materials): PO = 13.3 mg/kg, IV = 9.9 mg/kg, and IT = 0.18 mg/kg.

IT administration resulted in a C_max of_ 40.7 ± 11.7 ng/mL (mean ± SD) in whole blood, compared to PO administration with a C_max of_ 183.3 ± 48.6 ng/mL (p=0.88 compared to IT C_max)_ and IV administration with a C_max 1,_664.0 ± 724.6 ng/mL (p<0.005 compared to IT C_max,_ Supplementary Table 4). T_max in_ whole blood values for the IT, PO, and IV groups were 2-10 minutes, 4 hours, and 2-10 minutes, respectively. Mean (±SD) whole blood Area Under the Curve up to 24 hours post-dose (AUC_0-24hr)_ in the IT group was 89.3 ± 20.4 ng*hr/mL, compared with 2,320.3 ± 392.5 ng*hr/mL in the PO group (p<0.001) and 4,323.0 ± 331.5 ng*hr/mL in the IV group (p<0.001, Supplementary Table 4).

The different routes of administration were associated with notable differences in the HCQ concentrations in tissues and blood (Figure 2).

**Figure 2.**
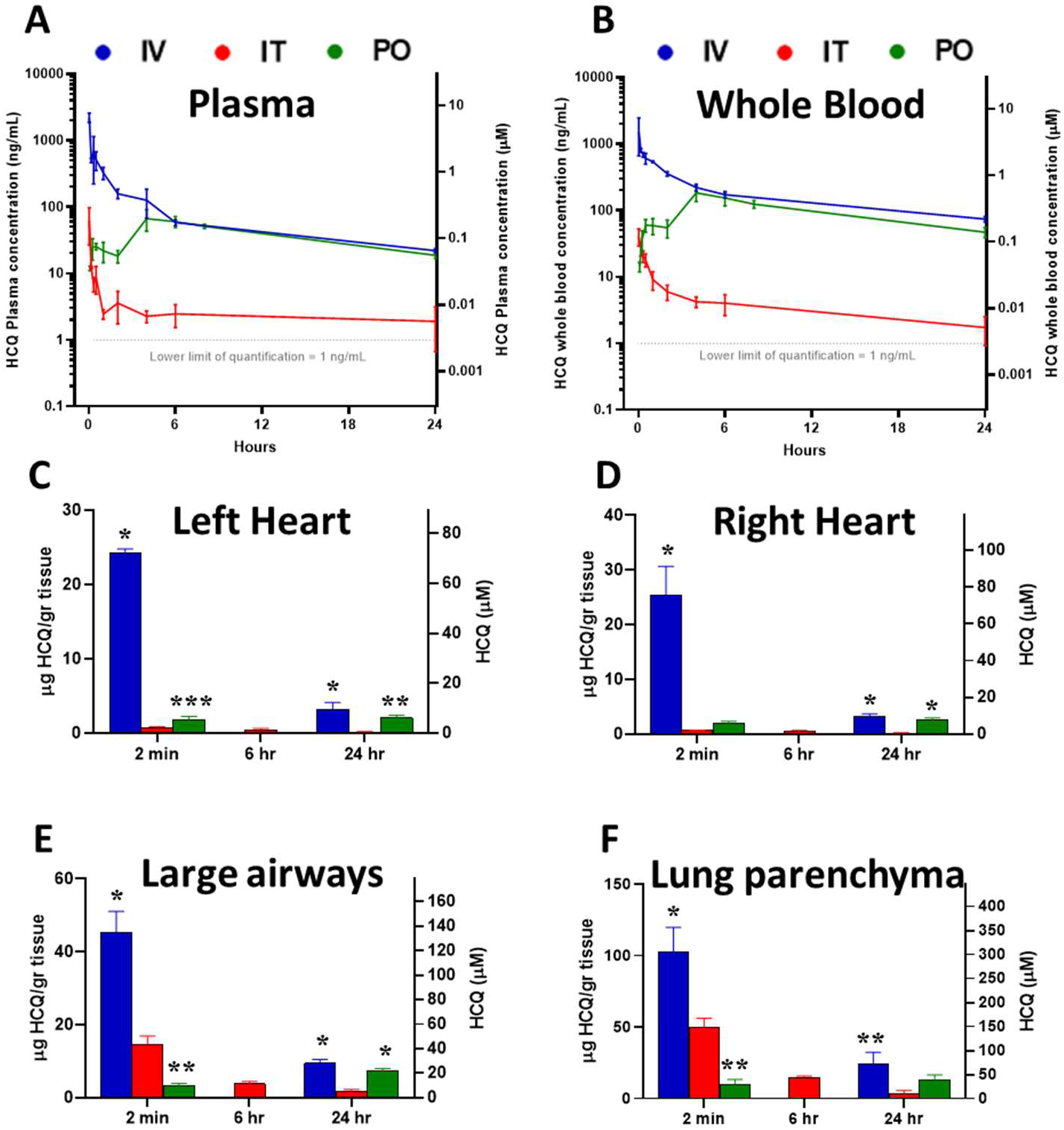
Concentration-time profiles of HCQ in plasma, whole blood, and tissue after single PO, IV, and IT administration of HCQS solution in male Sprague Dawley rats. Results at each timepoint are mean ± SD of measurements in 3 animals for each route of administration (IV 9.9 mg/kg, IT 0.18 mg/kg, PO 13.3 mg/kg), except for tissue concentrations at 6 hours, where they are the mean of values from 2 animals treated IT only. Results on y axes are scaled to both ng/mL or µg/gr (left axis) and µM (right axis). A) Plasma; B) Whole blood; C) Left heart; D) Right heart; E) Large airways; F) Lung parenchyma * p<0.001, ** p<0.01, *** p<0.05 compared to IT administration at the same timepoint

Thus, despite an IT dose that was ˂1.5% of the PO dose, at 2 minutes, IT administration was associated with higher HCQ concentrations in the large airways (14.61 ± 2.36 µg HCQ/g tissue, mean ± SD) and lung parenchyma (49.53 ± 6.51 µg/g) compared to PO administration (large airways: 3.19 ± 0.71, p<0.02; lung parenchyma: 9.88 ± 3.43, p<0.01). The large airway and lung concentrations decreased over 6 hours (decline of ∼75%) and 24 hours (decline of ∼90%) after IT administration, whereas these tissue concentrations increased by 24 hours after PO administration. By comparison, cardiac concentrations 2 minutes and 24 hours after IT administration were 60% and 93% lower than after PO administration (p<0.04 and p<0.01, respectively).

The ratios of the concentrations of HCQ in respiratory tract tissue (large airways and lung parenchyma) to either heart tissue (left or right heart), or blood (whole blood or plasma) were >1 at 2 minutes and at 24 hours post-dose with all three routes of administration (Table 1).

**Table 1.** Ratios of HCQ concentrations in large airways and lung parenchyma to left heart, right heart, whole blood, and plasma after single PO, IV, and IT administration of HCQS solution in male Sprague Dawley rats.

| Time | ROA | Large airways/ Left heart | Large airways/ Right heart | Large airways/ Whole blood | Large airways/ Plasma | Lung parenchyma / Left heart | Lung parenchyma/ Right Heart | Lung parenchyma/ Whole blood | Lung parenchyma / Plasma |
| --- | --- | --- | --- | --- | --- | --- | --- | --- | --- |
| 2 min | IT | 20.9 ± 5.1 | 24.4 ± 5.2 | 265.1 ± 51.7 | 377.0 ± 67.1 | 71.0 ± 16.4 | 82.8 ± 16.9 | 900.5 ± 173.2 | 1279.1 ± 204.7 |
|  | IV | 1.9 ± 0.2 | 1.8 ± 0.3 | 19.1 ± 4.0 | 20.5 ± 6.5 | 4.2 ± 0.7 | 4.0 ± 0.3 | 43.3 ± 11.5 | 46.2 ± 16.7 |
|  | PO | 1.8 ± 0.6 | 1.7 ± 0.6 | 45.5 ± 13.8 | 141.1 ± 25.3 | 5.5 ± 1.7 | 5.2 ± 1.7 | 141.9 ± 59.8 | 426.2 ± 75.2 |
|  | IT/PO* | 11.5 | 14.4 | 5.8 | 2.7 | 12.9 | 16.1 | 6.3 | 3.0 |
|  | p value (IT vs PO) | 0.022 | 0.016 | 0.013 | 0.016 | 0.019 | 0.015 | 0.010 | 0.011 |
| 24 hr | IT | 10.7 ± 1.4 | 10.2 ± 2.3 | 1025.6 ± 651.0 | 988.3 ± 597.8 | 24.4 ± 6.3 | 22.5 ± 1.7 | 2473.8 ± 1897.3 | 2033.1 ± 586.1 |
|  | IV | 3.1 ± 0.6 | 3.0 ± 0.1 | 128.5 ± 16.6 | 432.3 ± 71.2 | 8.0 ± 3.7 | 7.5 ± 2.1 | 333.4 ± 131.7 | 1119.5 ± 433.0 |
|  | PO | 3.6 ± 0.6 | 2.8 ± 0.4 | 155.6 ± 13.1 | 387.6 ± 3.4 | 6.6 ± 2.7 | 5.2 ± 2.0 | 282.7 ± 90.2 | 695.9 ± 182.8 |
|  | IT/PO* | 3.0 | 3.6 | 6.6 | 2.5 | 3.7 | 4.3 | 8.8 | 2.9 |
|  | p value (IT vs PO) | 0.006 | 0.028 | 0.147 | 0.224 | 0.025 | <0.001 | 0.183 | 0.048 |

\*The ratios are of means of HCQ concentration measurements in 3 animals for each route of administration and at each of the timepoints (total $n = 18$ ); $\mu\text{g HCQ/g tissue over } \mu\text{g HCQ/g tissue}$ or $\mu\text{g HCQ/g tissue over } \mu\text{g HCQ/mL blood or plasma}$ .

The ratios of respiratory tract tissue concentrations to heart tissue concentrations were, importantly, much higher after IT, compared to either IV or PO administration, with IT/PO ratios of 11.5-16.1 at 2 minutes (p<0.001) and 3.0-4.3 at 24 hours post-dose (p<0.005). At 6 hours post-dose, tissue concentrations were only available in rats that were administered HCQS by the IT route (Figure 2 C,D,E,F): ratios of respiratory tract tissue concentrations to heart tissue concentrations were 9.3-39.0 and ratios of respiratory tract tissue concentrations to blood (whole blood or plasma) were 771.0-3,186.6. As HCQ doses were adjusted to reflect human doses, these results would suggest that following single doses, administering HCQ directly to the respiratory tract may lead to higher airway and lung exposure without exposing the heart and other tissues to HCQ concentrations that are above those achieved via oral dosing.

### Human Phase 1 study

#### Enrollment and demographics

Between June and August 2020, 12 volunteers were screened, of whom 10 were enrolled into the study (Figure 3).

**Figure 3.**
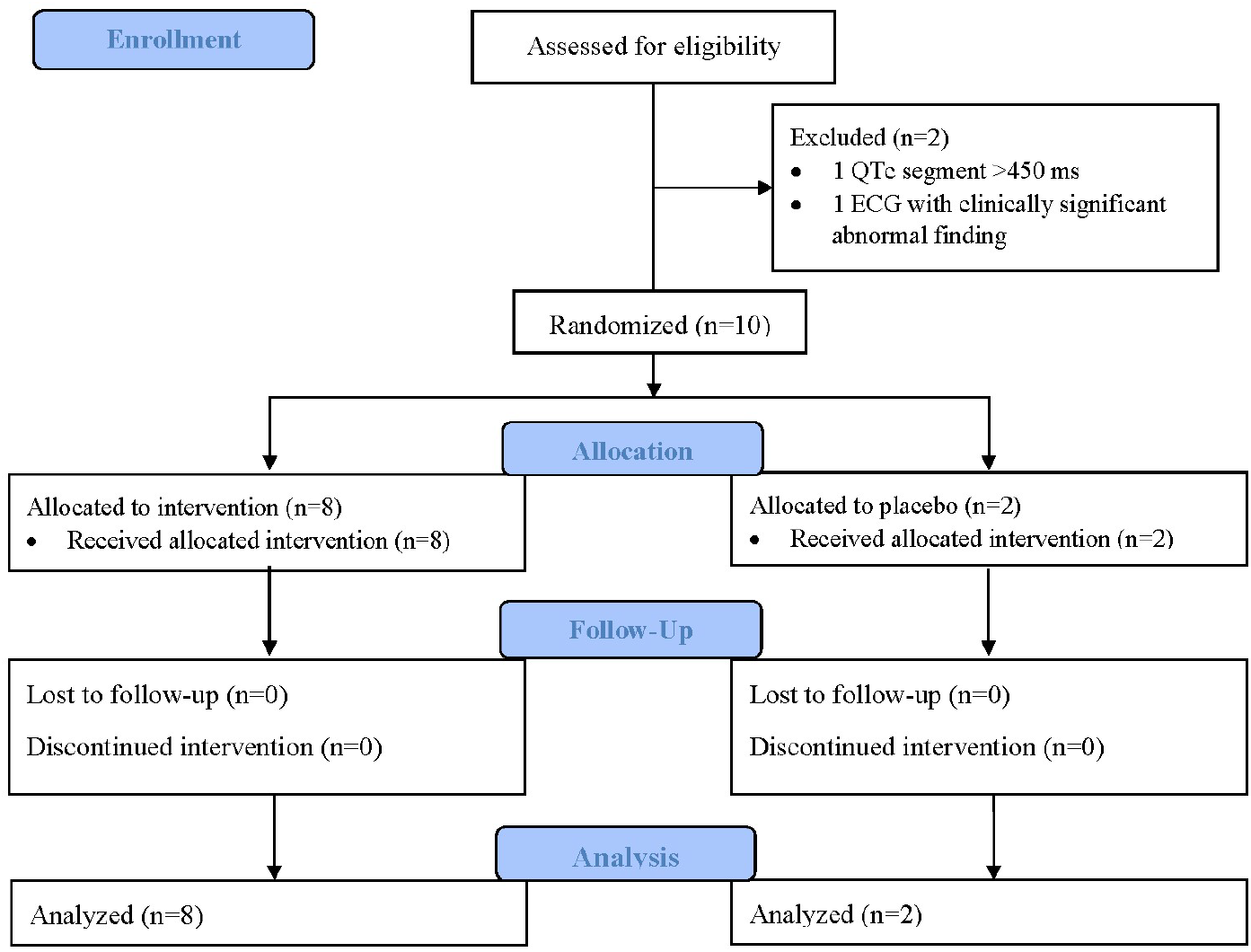
Phase 1 Trial profile.

Two sentinel participants were initially enrolled into the aHCQS 20 mg dose cohort in a single-blind manner. After review of the results from these 2 participants, the safety review committee recommended enrolling 2 sentinel participants into the aHCQS 50 mg dose cohort. After another safety review of these 2 participants, the safety review committee recommended randomization of 6 additional participants in that cohort to aHCQS or placebo (4:2) (Figure 4).

**Figure 4.**
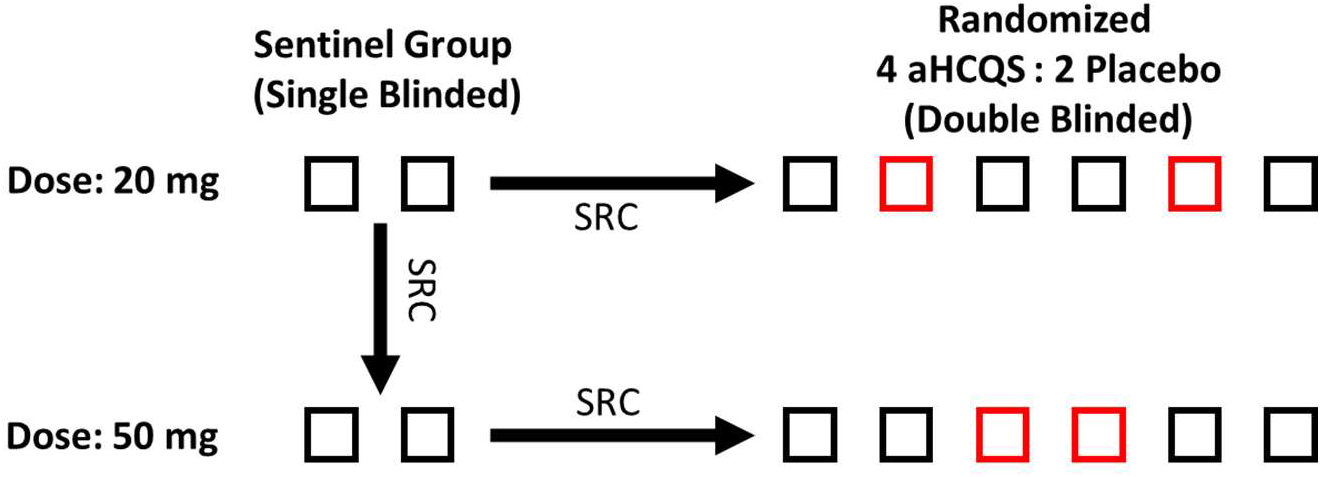
Original aHCQS phase 1 dose-escalation study design. Note that after first 2 sentinel participants received the 20 mg dose, the safety review committee (SRC) recommended advancing to the next dose level, and proceeding to the 2 sentinel participants receiving 50 mg rather than the randomized group at 20 mg. As a result, a total of 10 participants were tested rather than the originally proposed 16. Red boxes signify randomization to placebo and black boxes signify randomization to aHCQS.

The baseline characteristics and clinical demographics by cohort are shown in Table 2.

**Table 2.**
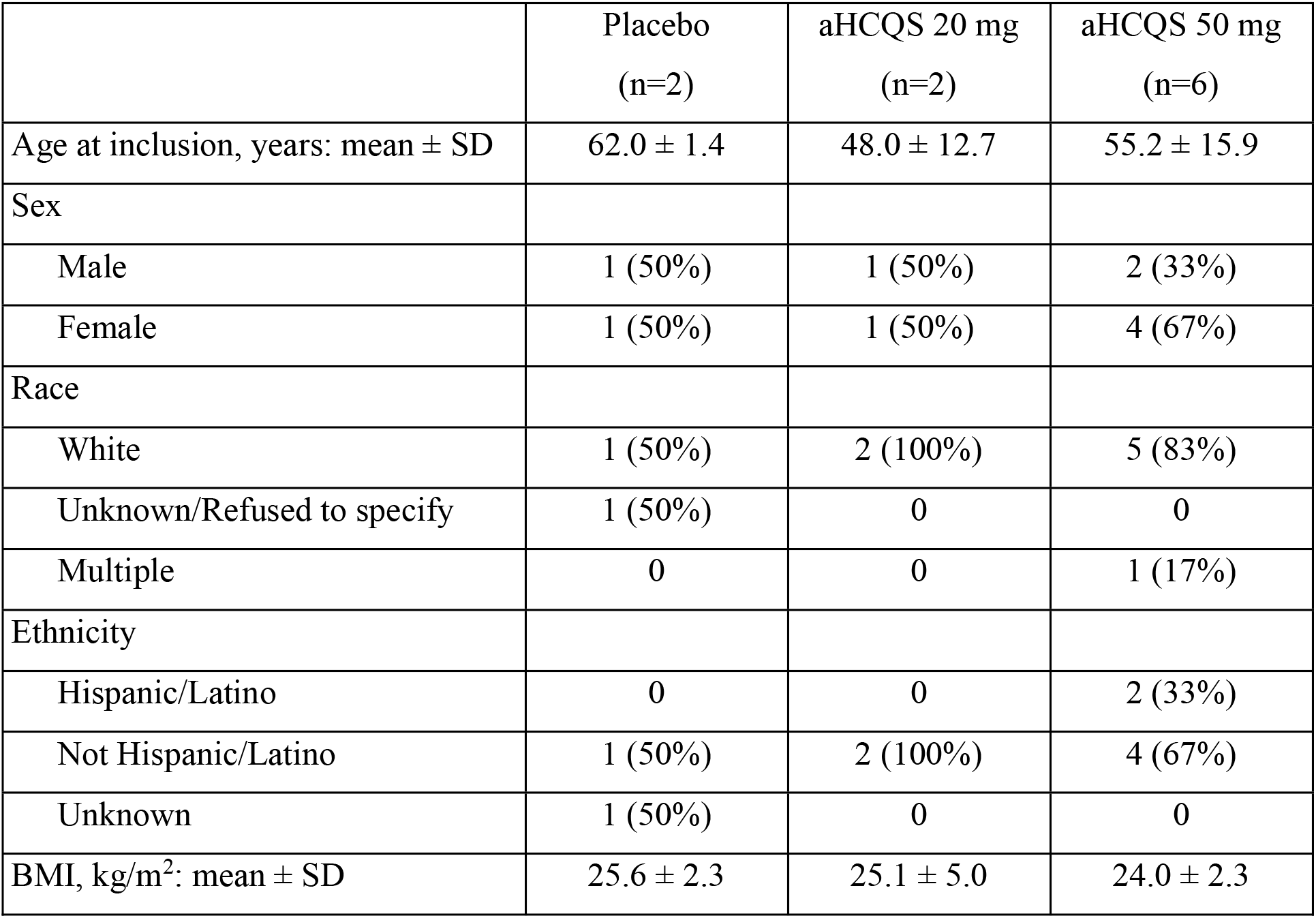
Demographics and baseline characteristics of participants.

The majority of participants (6/10) were women, and the mean (±SD) age was 55 ± 13 years.

#### Study drug administration

All participants completed the inhalation during the first attempt and none of the participants required a pause during the inhalation. All the participants were observed to inhale the study drug through the mouthpiece and exhale through the nose. Despite equivalent volumes, but in accord with data from the breath simulator (see Supplementary Methods) and published data (38), solute concentration affected the nebulization time. Thus, the mean (±SD) total inhalation time in the aHCQS 50 mg cohort (245 ± 19 seconds) was significantly longer than in the aHCQS 20 mg cohort (141 ± 16 seconds, p<0.001) or in the placebo cohort (129 ± 18 seconds, p<0.001).

### Tolerability assessments

#### Adverse events

No serious adverse events (SAEs) were observed. All adverse events (AEs) were graded mild in severity (Table 3) and were transient.

**Table 3.** Summary of treatment-emergent adverse events.

|  | Placebo<br>(N=2)<br>n (%) [E] | aHCQS 20 mg<br>(N=2)<br>n (%) [E] | aHCQS 50 mg<br>(N=6)<br>n (%) [E] |
| --- | --- | --- | --- |
| Number of participants with at least one treatment-emergent adverse event | 0 | 2 (100%) [3] | 4 (67%) [6] |
| Blood bilirubin increase, grade 1 | 0 | 0 | 1 (17%) [1] |
| Dizziness, grade 1 | 0 | 1 (50%) [1] | 2 (33%) [2] |
| Dysgeusia, grade 1 | 0 | 2 (100%) [2] | 3 (50%) [3] |

N = number of participants dosed for each treatment; n = number of participants reporting at least one incidence of the specified adverse event; [E] = number of events

No AEs were observed in the placebo arm. In total, 9 treatment-emergent AEs were observed in 6/8 participants who received aHCQS [2/2 in the 20 mg cohort (100%) and 4/6 in the 50 mg cohort (67%)]: 5 cases of transient dysgeusia lasting <90 minutes after completion of the inhalation, 3 cases of dizziness lasting <60 minutes after completion of the inhalation, and a single case of increased bilirubin (total 1.9 mg/dL, direct 0.5 mg/dL) on Day 8 that resolved on repeat testing performed 1 month after administration of the study drug. All cases of dysgeusia were judged to be related to the study drug.

#### Electrocardiographic assessments

ECGs were performed at screening, pre-dose, and at 2 and 6 hours and 1 and 7 days post-inhalation. QT segment durations, corrected with the Bazett formula (QTc), were minimally changed from baseline (Figure 5A) after 1-6 hours, and after 1 and 7 days; all were ≤455 ms.

**Figure 5.**
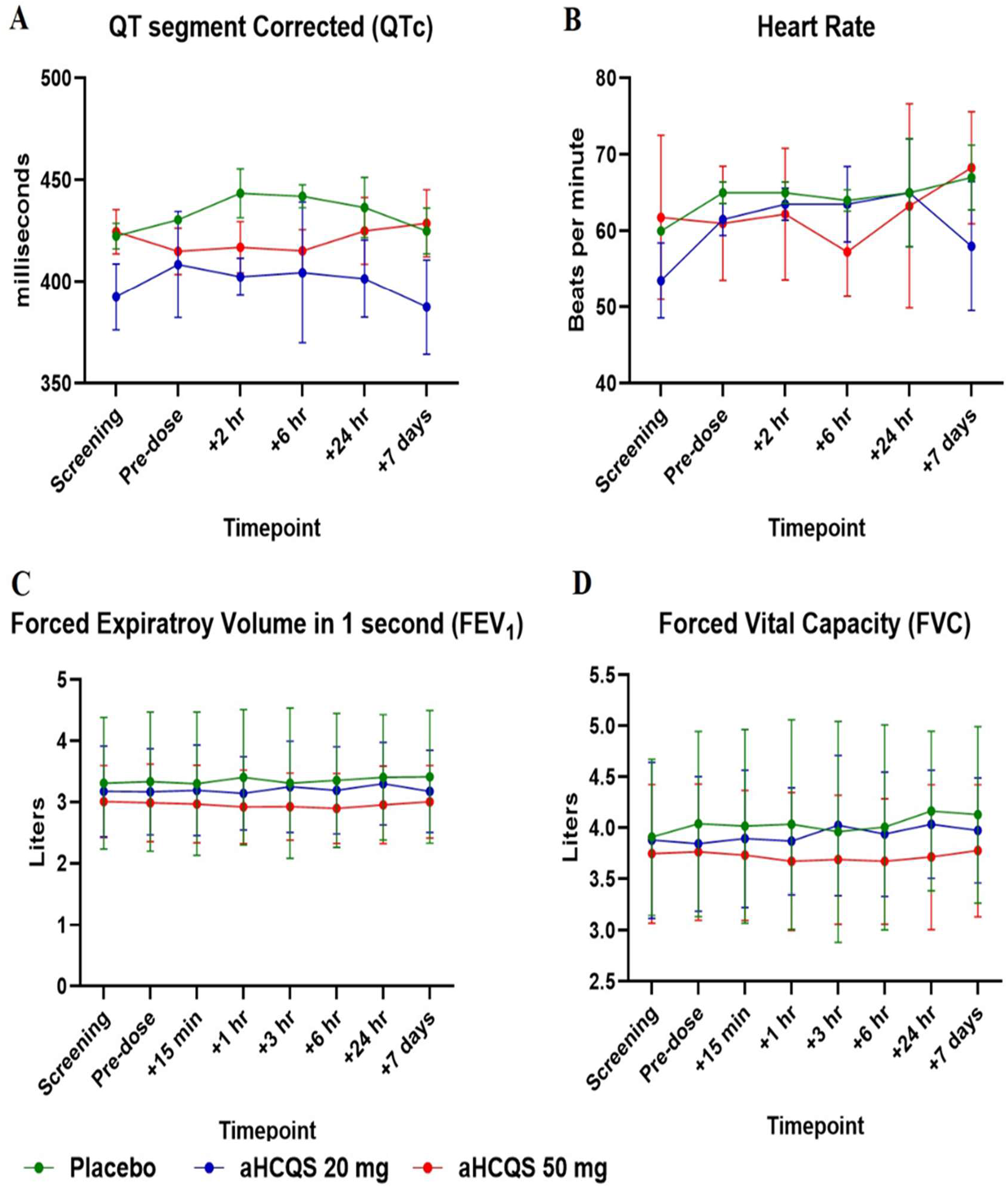
ECG and PFT data. ECG and PFT study assessments after inhalation of placebo (*n* = 2) or aHCQS (20 mg, *n* = 2; 50 mg, *n* = 6). Results shown are mean ± SD. A) Corrected QT segment; B) Heart rate; C) Forced Expiratory Volume in 1 second (FEV_1);_D) Forced Vital Capacity (FVC).

The maximum prolongation in QTc recorded during the study was 34 ms, identified as an isolated finding on the Day 8 ECG of a participant in the aHCQS 50 mg cohort (screening: 423 ms; pre-dose: 414 ms; +2 hours: 420 ms; +6 hours: 402 ms; Day 2: 420 ms; Day 8: 448 ms). Heart rate was minimally changed throughout the study (Figure 5B).

#### PFT assessments

FEV_1 an_d FVC were unchanged from baseline after 15- 360 minutes and after 1 and 7 days (Figure 5 C,D).

#### Participant-reported outcomes

Based upon questionnaire results:

##### Sensory measures

The participants rated the smell, taste, and bitterness of the study drug on 6-level Likert scales. Participants in the placebo and active study drug arms rated the smell of the drug similarly as either “no smell” or “slight smell.” Participants who received placebo rated the study drug’s taste as “no taste” or “slight taste, not unpleasant.” Most of the participants who received aHCQS found the study drug to have an unpleasant bitter taste, but all rated both the taste and bitterness as tolerable (Supplementary Table 8).

##### Acceptability

In response to questions about willingness to use the study medication on a regular basis, all participants who received the active study drug responded that they would be willing to use it on a regular basis and that the taste of the study medication would not keep them from using it on a regular basis. Even though none of the participants had extensive experience with using a nebulizer, 90% reported the use of the Aerogen inhalation system as “very easy” (40%) or “easy” (50%); a single participant (10%) reported that using the inhalation system was “somewhat difficult” (Supplementary Table 9).

##### Exhalation through the nose

In response to the question “during the inhalation of the study drug, were you able to exhale through your nose as you were encouraged?,” 60% of participants responded “all of the time,” 30% responded “most of the time,” and a single participant (10%), who was enrolled in the placebo arm, responded “some of the time” (Supplementary Table 10).

##### Cough

None of the participants in the placebo arm reported developing a cough within 3 hours after completing the inhalation. In the aHCQS 20 mg cohort, one participant reported that they had cough “none of the time” and the other reported “hardly any of the time.” In the AHCQS 50 mg cohort, 2/6 (33%) replied “none of the time,” 2/6 (33%) replied “hardly any of the time,” a single participant (17%) replied “a little of the time,” and a single participant (17%) replied “some of the time.” On Days 2 and 8, all of the participants in the placebo and aHCQS 20 mg cohorts responded that they did not suffer from cough since completing the Day 1 questionnaire. In the aHCQS 50 mg cohort, 4/6 (67%) responded that they had no cough after completing the Day 1 questionnaire, and 2/6 (33%) responded that they coughed “a little of the time” or “hardly any of the time.” The participants graded their cough severity on a visual analog scale (VAS) with 0 representing “no cough” and 100 representing “worst cough ever.” In the aHCQS 50 mg cohort, the mean (±SD) VAS score was 17.5 ± 9.6 in the 4/6 who replied that they had cough on Day 1, and 10.0 ± 0 in the 2/6 who replied that they had cough on days 2-8 (Supplementary Table 11).

#### Pharmacokinetic Data

Mean concentration-time profiles of HCQ in whole blood and plasma at each dose level are shown in Figure 6 and key PK parameters are summarized in Table 4.

**Figure 6.**
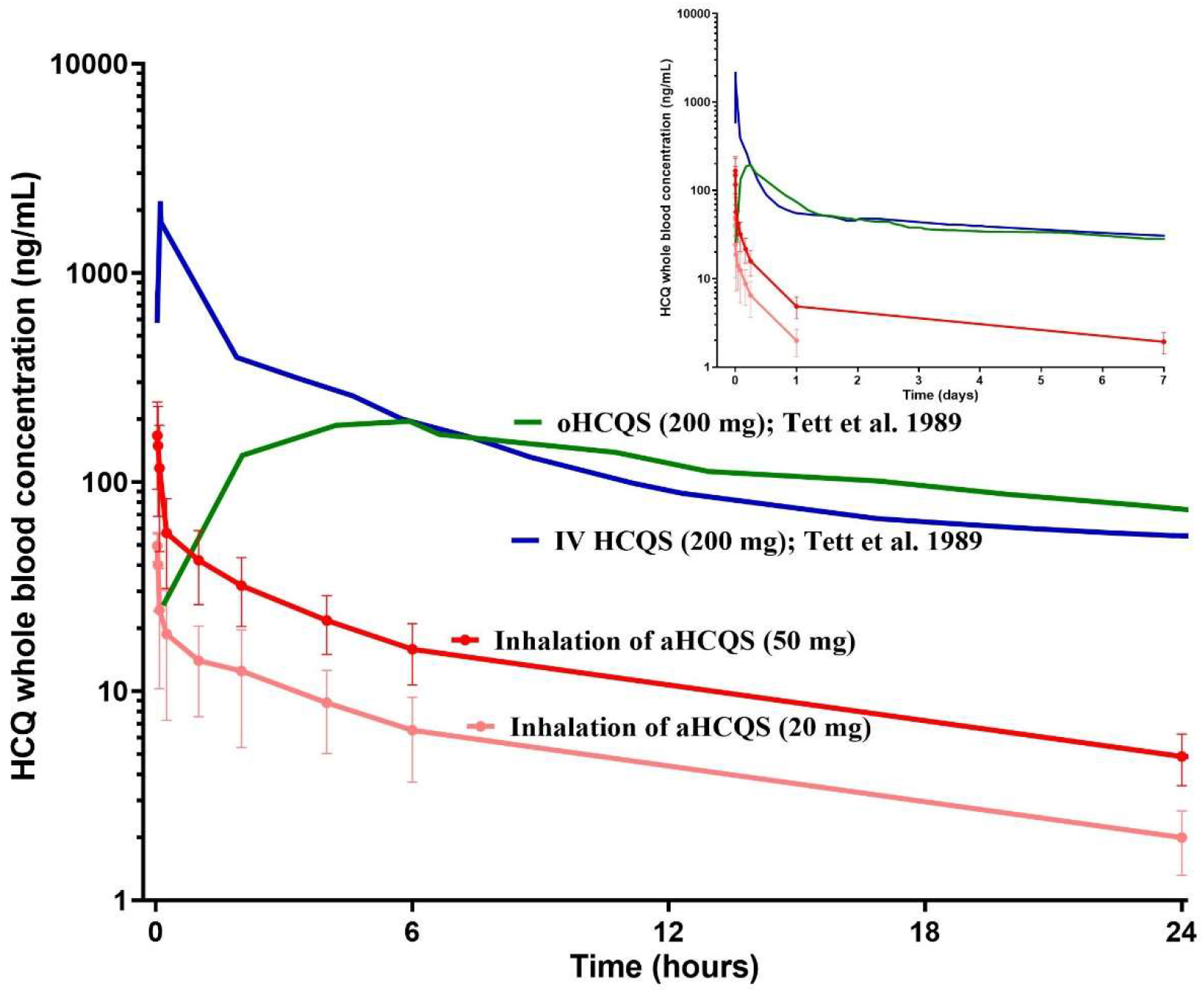
Mean concentration-time profiles of HCQ in whole blood after administration of aHCQS in comparison to published data on HCQS administered IV and PO. Results shown for aHCQS (aHCQS 20 mg, *n* = 2; aHCQS 50 mg, *n* = 6) are mean ± SD on a log scale. The last quantifiable concentration after aHCQS administration was 24 hours in the aHCQS 20 mg cohort and 7 days in the aHCQS 50 mg cohort. HCQ concentrations after oHCQS and IV HCQS were extracted from figures by Tett et al., 1989 (7) using WebPlotDigitizer. The inset shows the full timescale (0-7 days), and the large figure shows an expanded time scale (0-24 hours).

**Table 4.**
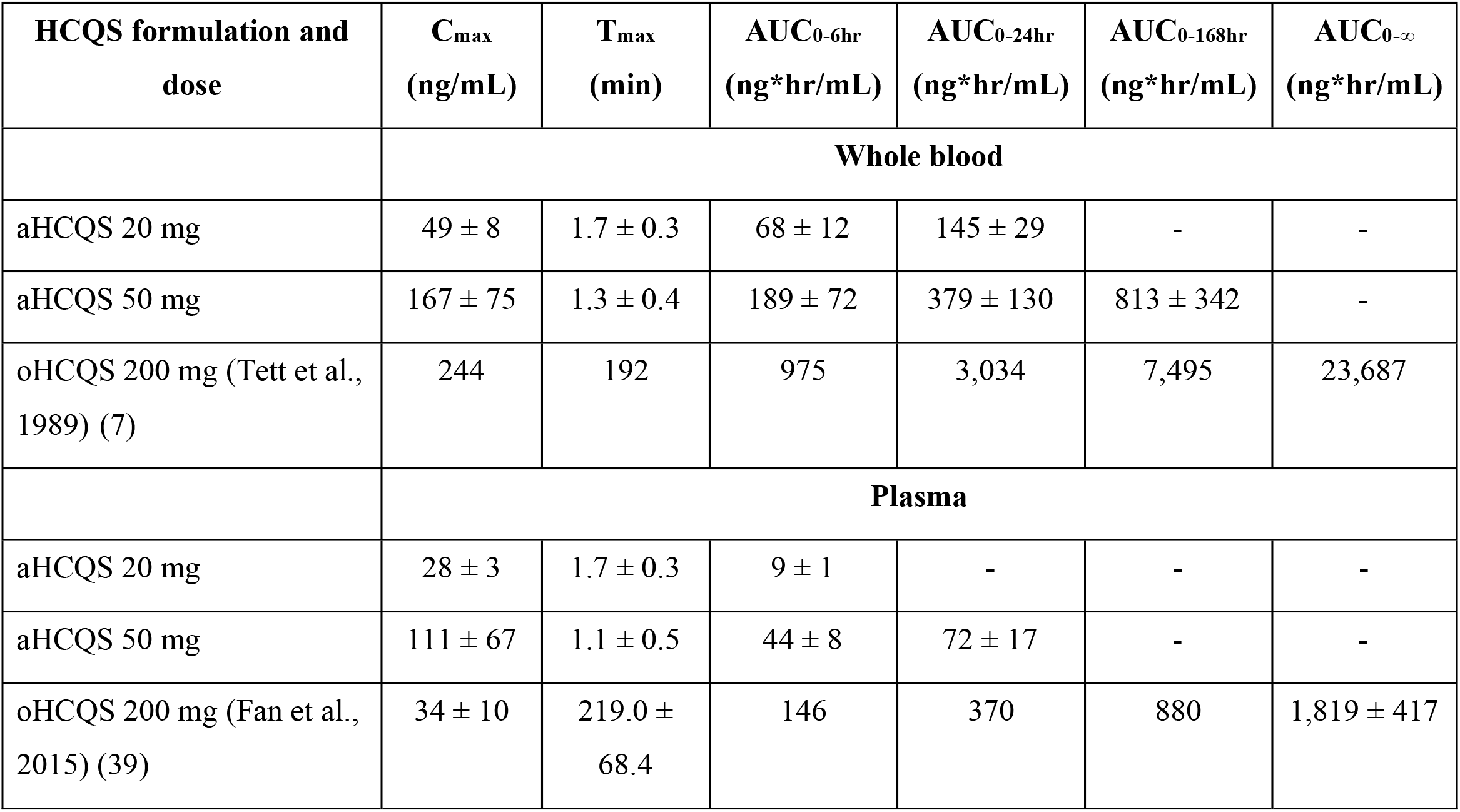
Pharmacokinetic parameters in the Phase 1 study of aHCQS in comparison to published data on PO HCQS. Results shown for aHCQS (aHCQS 20 mg, *n* = 2; aHCQS 50 mg, *n* = 6) are mean ± SD. AUC_0-24hr an_d AUC_0-168hr af_ter aHCQS are only provided if HCQ was quantifiable at the indicated final timepoint (lower limit of detection: 1 ng/mL in whole blood, 0.477 ng/mL in plasma). AUC_0-∞ is_not provided for aHCQS because it cannot be determined reliably based on the data (the terminal phase of HCQ PK could not be adequately characterized because the number of samples after 24 hours was not sufficient to produce a terminal log-linear phase). HCQ concentrations in whole blood and plasma after oHCQS were reported by Tett et al., 1989 (7) and Fan et al., 2015 (39). AUC in whole blood after oHCQS was calculated with the formula provided in Tett et al. and AUC in plasma after oHCQS was extracted from figures in Fan et al. using WebPlotDigitizer. Doses in the left column are of HCQS, and all concentrations in the other columns are of hydroxychloroquine base.

| HCQS formulation and dose | C<sub>max</sub><br>(ng/mL) | T<sub>max</sub><br>(min) | AUC<sub>0-6hr</sub><br>(ng*hr/mL) | AUC<sub>0-24hr</sub><br>(ng*hr/mL) | AUC<sub>0-168hr</sub><br>(ng*hr/mL) | AUC<sub>0-∞</sub><br>(ng*hr/mL) |
| --- | --- | --- | --- | --- | --- | --- |
|  | **Whole blood** |  |  |  |  |  |
| aHCQS 20 mg | 49 ± 8 | 1.7 ± 0.3 | 68 ± 12 | 145 ± 29 | - | - |
| aHCQS 50 mg | 167 ± 75 | 1.3 ± 0.4 | 189 ± 72 | 379 ± 130 | 813 ± 342 | - |
| oHCQS 200 mg (Tett et al., 1989) (7) | 244 | 192 | 975 | 3,034 | 7,495 | 23,687 |
|  | **Plasma** |  |  |  |  |  |
| aHCQS 20 mg | 28 ± 3 | 1.7 ± 0.3 | 9 ± 1 | - | - | - |
| aHCQS 50 mg | 111 ± 67 | 1.1 ± 0.5 | 44 ± 8 | 72 ± 17 | - | - |
| oHCQS 200 mg (Fan et al., 2015) (39) | 34 ± 10 | 219.0 ± 68.4 | 146 | 370 | 880 | 1,819 ± 417 |

aHCQS was absorbed rapidly, with C_max ge_nerally observed in whole blood and plasma 2 minutes after completion of the inhalation. Thereafter, HCQ levels in whole blood and plasma declined in a multi-exponential manner, with rapid distribution in the 15 minutes after the end of inhalation. Whole blood HCQ levels were detectable at concentrations ≥1 ng/mL for up to 24 hours in all participants in the aHCQS 20 mg cohort and for up to 168 hours in 5/6 participants in the aHCQS 50 mg cohort. Plasma HCQ levels were detectable at concentrations ≥0.477 ng/mL for up to 4-6 hours in the participants from the aHCQS 20 mg cohort and up to 24 hours in 4/6 participants from the AHCQS 50 mg cohort.

The ratio of HCQ whole blood to plasma concentrations was highly variable at low concentrations, and less so at values >70 ng/mL (overall mean ± SD: 5.6 ± 4.8, range: 1.4-26.5; at concentrations >70 ng/mL, 1.7 ± 0.4, range: 1.4-3.0; Supplementary Figure 2A). Of the 80 post-dose samples that were collected and analyzed, HCQ concentrations were below the limit of detection in both whole blood and plasma in just 3 samples, all on Day 8; in 10 samples HCQ was detected in whole blood (concentration range: 1.21-8.51 ng/mL) but not plasma.

Comparison of PK characteristics between cohorts showed that HCQ C_max va_lues increased in a greater than dose proportional manner, increasing 3.4-fold in whole blood and 4-fold in plasma, for a 2.5-fold increase in dose. However, the increase in AUC_0-24hr in_ whole blood was dose-proportional, increasing 2.6-fold for a 2.5-fold increase in dose.

#### Estimation of respiratory tissue HCQ concentration

The initial estimated regional (extra-thoracic, bronchial, bronchiolar, alveolar-interstitial) concentrations of HCQ in the ELF achieved immediately after a single inhalation of aHCQS at a dose of 20 mg or 50 mg were >2,000 µM, a value that surpasses the highest half-maximal inhibitory concentration (IC_50)_ of HCQ required to inhibit SARS-CoV-2 replication in the reported in vitro experiments (119 µM) (40). The time course of the subsequent drop in ELF concentration due to absorption depends on airway epithelial permeability, which is not well-characterized and thus not amenable to modeling.

Estimation of tissue concentrations after administration of aHCQS is more speculative because there are no available direct measurements of human HCQ tissue:blood ratios as a function of time. Thus, the respiratory tissue concentrations were estimated based on extrapolations from the observed tissue:blood HCQ concentration ratios at different timepoints in rats (Supplementary Table 12). The HCQ concentrations in the large airways and the lung parenchyma 2 minutes post-dose and in the lung parenchyma 6 hours post-dose were estimated to exceed the IC_50 va_lue for inhibiting SARS-CoV-2 replication after a single dose of aHCQS 50 mg, but the concentrations decrease dramatically by 24 hours (Supplementary Table 12).

### Impact of IT and IN HCQS solution on the Golden Syrian hamster preclinical model of SARS-CoV-2 infection

Nineteen animals were studied, with 5 in the control group, 7 receiving IT HCQS solution at 0.036 mg, equivalent to a human dose of 60 mg loaded in the nebulizer (low dose group), and 7 receiving HCQS solution at 0.072 mg, equivalent to a human dose of 120 mg loaded in the nebulizer (high dose group) (Supplementary Table 3). HCQS solution was administered IT daily on days -3, -2, and -1; the SARS-CoV-2 challenge was performed on day 0; and IN HCQS solution was administered 4 hours post-challenge (low dose: 0.12 mg, high dose 0.24 mg).

There were no unscheduled deaths. Reductions in body weight, an indicator of SARS-CoV-2 infection, occurred to the same extent in all three groups, reaching ∼13- 15% by day 7 (Figure 7A).

**Figure 7.**
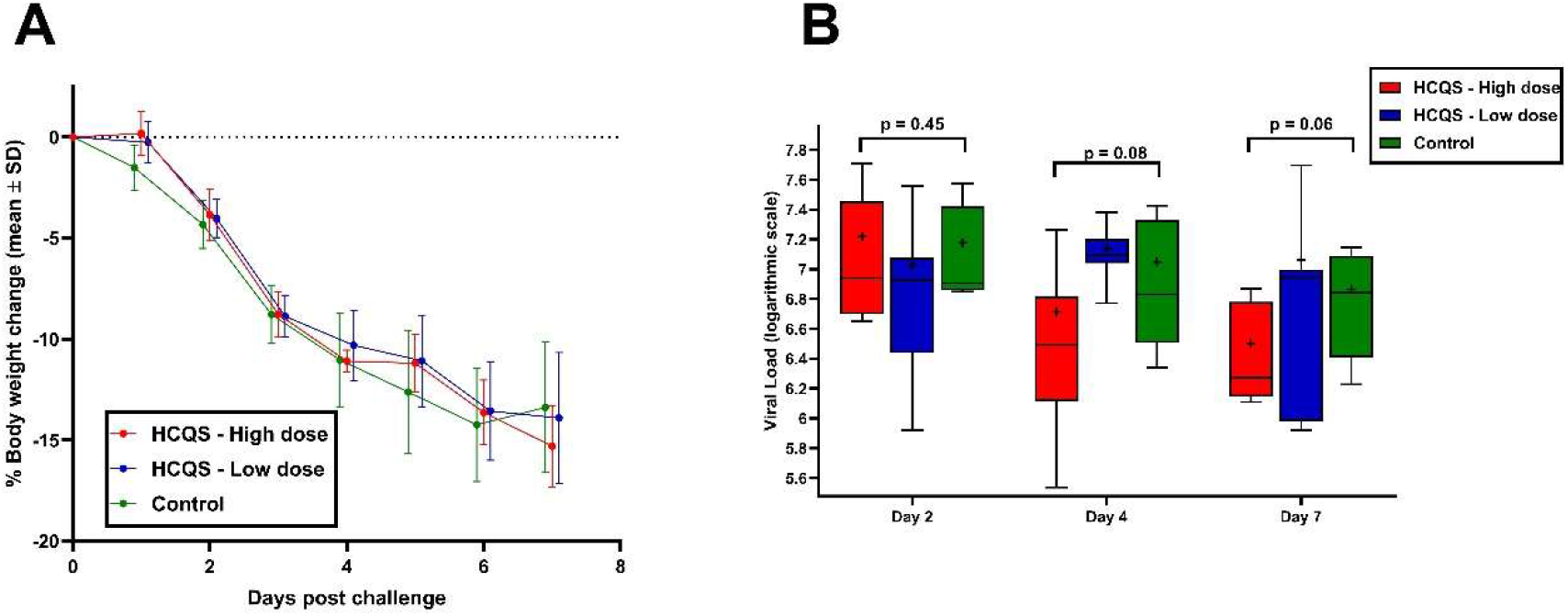
Impact of intratracheal (IT) and intranasal (IN) HCQS on the Golden Syrian hamster preclinical model of SARS-CoV-2 infection. Golden Syrian hamsters were treated with IT HCQS on days -3, -2, and -1, followed by SARS-CoV-2 challenge, and IN HCQS 4 hours post-challenge. A) Percent body weight change post-challenge; B) SARS-CoV-2 Viral loads in oral swabs on days 2, 4, and 7 post-challenge (horizontal line = median, + = mean, box = interquartile range, whiskers = range, p-values provided for high dose vs control group)

SARS-CoV-2 viral load measurements were also similar among the three groups on days 2, 4, and 7, although there was a trend for a reduction in SARS-CoV-2 viral load in the high dose group compared to the other two groups on days 4 and 7 that was not statistically significant (Figure 7B; p-values for comparison of viral load between the high-dose group and the control group were 0.45 on Day 2, 0.08 on Day 4, and 0.06 on Day 7, p-values for comparison of viral load between the low-dose group and the control group were 0.14 on Day 2, 0.83 on day 4, and 0.39 on day 7). At scheduled necropsy on day 7, the lungs of all 19 animals were found on gross examination to be hemorrhagic, and microscopic examination confirmed the presence of hemorrhage. Lung lesions consistent with SARS-CoV-2 infection (34, 41) were found in animals from all study groups, and included a range of inflammatory and reactive epithelial changes in small bronchi, terminal respiratory bronchioles, and adjacent alveoli (Supplementary Table 13, Supplementary Figure 4). Microscopic lesions affected approximately 50-70% of the total lung area visible in histologic sections from most animals. No differences in the incidence or severity of microscopic findings were evident across study groups.

## Discussion

The profound global impact of the COVID-19 pandemic elicited a monumental effort to identify effective therapies. Early in vitro studies demonstrating activity of HCQS in inhibiting viral replication, reinforced by previous in vitro studies showing antiviral activity of HCQS against the related SARS-CoV virus (8, 10, 40, 42–47), and anecdotal human studies suggesting more rapid clearance of virus in patients treated with oHCQS (11) led to the registration of 160 studies estimated to include over 250,000 participants by November 2020 (48). Unfortunately, these studies utilized primarily oral HCQS administration, thus leaving open the possibility that the lack of drug effect was due to either slow onset of drug action due to the large volume of distribution or insufficient drug concentrations in target tissues.

Three major limitations in our knowledge at the beginning of the pandemic compromised developing a rational approach to assessing the potential utility of aHCQS:

**1. Extrapolating from in vitro assays using cultured cell lines to in vivo human respiratory tract and lung tissue.** The in vitro studies employed a variety of cell lines obtained from different animals and different organs. Most importantly, while multiple early studies using an African green monkey kidney epithelial cell line (Vero E6) and a single study using a human hepatoma cell line (Huh-7) reported IC_50 va_lues of HCQS from 1 to 13 µM (8, 10, 42–47), a later study using the Calu-3 cell line, which is thought to be more reflective of lung epithelial cells, reported an IC_50 of_119 µM (40).
**2. Sparse availability of PK data on HCQS.** Despite its widespread use in rheumatologic disorders, at the beginning of the pandemic, there were only limited published data on the PK of HCQS via all routes of administration. These studies indicated that for oHCQS bioavailability is ∼0.75, its volume of distribution is large, and it has a very long terminal half-life, implying sequestration by, and slow release from, tissues (7, 12, 20, 39, 49–51). The latter was supported by reports of higher whole blood than plasma concentrations (7).
**3. Limited understanding of the mechanism by which HCQS inhibits viral replication and impacts COVID-19.** Proposed mechanisms by which HCQS inhibits SAR-CoV-2 replication include: a. Increasing endosomal pH in target cells, leading to, among other effects, reduced viral entry, replication, and/or budding (e.g., membrane fusion, acid-dependent proteolytic cleavage of the virus’ S protein, post-translational modification of envelope glycoproteins). b. Inhibiting glycosylation of ACE2, the cell surface viral receptor. One would expect the first mechanism to correlate with the concentration of HCQ inside the cell, or more specifically in the cell’s endosomes, whereas the second mechanism is likely to depend on the concentration of the fluid surrounding the cell. Since the reported tissue culture experiments were conducted by bathing the cells in buffer containing the drug, but without differentiating tissue versus medium concentrations, they did not provide data to assess the relative contributions of drug in each compartment.

Later data indicated that SARS-CoV-2 gains entry into cells after the spike (S) protein engages the ACE2 receptor either by cleavage of the S protein at the S2’ site at the cell membrane by the serine protease TMPRSS2 or cleavage by cathepsin L after clathrin-mediated endocytosis into endolysosomes (Figure 1) (52–55). HCQ inhibition of endolysososomal acidification is only expected to affect the cathepsin L mechanism, which probably accounts for the reported very weak effects on SARS-CoV-2 replication in nasal goblet secretory cells and type II pneumocytes, which express both ACE2 and TMPRSS2, as well as its very weak effects on SARS-CoV-2 infection. In support of this hypothesis, experimental evidence indicates that combined inhibition of both TMPRSS2 and cathepsin L is much more effective at preventing SARS-CoV-2 entry than either inhibitor alone (53). This raises the possibility that combining a TMPRSS2 inhibitor with HCQ may be effective at ameliorating SAR-CoV-2 infection. A recent double blind clinical study found that the TMPRSS2 inhibitor camostat did not significantly improve clinical outcomes in patients with COVID-19 (54), suggesting that there may well be a role for simultaneous inhibition of both pathways. If that is borne out, aHCQS may well be an attractive addition to TMPRSS2 inhibition.

The ultimate effects of HCQ on the innate immune response to SARS-CoV-2 remain unknown but may also be important to consider as they may modulate the inflammatory phase of COVID-19 (56). HCQ has been reported to reduce Toll-like receptor 7 (TLR7) affinity for viral RNA via endosomal alkalization, resulting in reduction of cytokine induction, and to inhibit cyclic GMP-AMP synthase (cGAS) activity in host cells, thereby leading to decreased type I interferon (IFNβ) production (55). However, characterizing the PK and pharmacodynamics of HCQ’s anti-inflammatory effects is even more complex than characterizing the antiviral effects.

To begin to address the uncertainties noted above, we conducted an integrated series of studies to critically assess the potential for aHCQS to achieve respiratory tract concentrations adequate to inhibit SAR-CoV-2 replication without producing toxic cardiac effects:

### 1. Preclinical PK study to compare HCQ tissue distribution after single PO, IV, and IT administration of HCQS solution in rats

Although the IT HCQS dose was less than 1.5% of the oral dose (0.18 mg/kg vs 13.3 mg/kg), it achieved immediate respiratory tissue concentrations that were >4-fold higher than those observed after PO administration. At the same time, cardiac tissue levels were much lower, which might translate into a lower risk of QT segment prolongation in humans. Thus, these experiments supported the hypothesis that administering HCQS directly to the respiratory tract may achieve a much more favorable distribution between the target tissues in the respiratory tract versus the heart soon after administration.

However, there was no evidence that HCQ immediately gets sequestered in the respiratory tissue since there was a rapid, albeit short-lived, increase in blood levels (T_max 2-_10 minutes) after single dose administration, and lung tissue levels dropped by ∼70% within 6 hours, and ∼90% by 24 hours. Nonetheless, lung tissue:blood HCQ concentration ratios were >250 at both the 2 minute and 24 hour sampling point, suggesting some retention of HCQ in the lung. Similar findings have been reported in other animal studies. In macaques treated with oHCQS, the decline in blood HCQ concentrations over time was faster than the concomitant decline in lung tissue HCQ concentrations (57), and in rats receiving IT HCQ, lung HCQ concentrations remained elevated over a 72 hour period (58). Of note, in the latter study, the earliest timepoint for sampling blood/tissue was 15 minutes post-dose. Based on our data and the published data on HCQ PK (7), collection of samples at earlier timepoints is crucial for characterizing the PK of HCQ after IT or IV administration, since the C_max,_ measured at 2-3 minutes, is several fold higher than the concentrations measured at 15 minutes.

Collectively, these experiments demonstrate that compared to oral administration, aHCQS may have a more favorable therapeutic index when considering respiratory tract, blood, and heart exposure, especially as the heart is a site where serious side effects have been described.

### 2. Phase 1 tolerability and PK study in humans

We found that aHCQS delivered with the Aerogen inhalation system was well-tolerated by study participants, with all of them completing the inhalation in a single attempt and being able to exhale through their nose. The bitter taste was acceptable and there was minimal cough. There was an initial short-lived blood peak of HCQ, which contrasts with the previously reported slower rise in blood HCQ after oral administration (7). The early peak in blood levels 2 minutes after completing the inhalation in our Phase 1 study, however, contributed only a minor fraction of the total AUC at 168 hours (AUC_0-1hr:_ 62 ng*hr/mL vs AUC_0-168hr:_ 813 ng*hr/mL). As with oral administration, there is a long terminal phase component after aerosolized administration (7). The latter is thought to reflect tight tissue binding of HCQ, but it is unclear if binding or “tissue drug loading” is enhanced in the organs and tissue the drug first encounters (e.g., respiratory tract via aerosol inhalation and intestines via oral administration). The data from the rat study and our Phase 1 study indicate that HCQ is initially taken up by respiratory tissues, but at the gross organ level it does not appear to remain in the large airways or lung parenchyma for a protracted period of time after just a single dose of aHCQS. Our studies do not provide direct evidence as to whether it is retained in target cells or organelles or accumulates intracellularly after multiple doses.

### 3. Assessment of efficacy of IT/IN HCQS in a SARS-CoV-2 hamster model

To gain additional insights into the potential utility of aHCQS in treating COVID-19 we utilized the Syrian hamster model because a consensus emerged as to its utility in assessing interventions to prevent or treat SARS-CoV-2 infection (36). To minimize the likelihood of missing a beneficial effect of aHCQS, we pretreated the animals IT for three days before the viral challenge, and treated them once IN after the challenge. Despite this, there was no evidence of protection against infection based on animal weight loss or pulmonary pathology. Viral load assessments did show a trend toward lower viral loads with the high dose of HCQS that was not statistically significant, but since this was not associated with clinical or pathological amelioration of infection, it is of uncertain significance.

Extensive modelling of the PK of pulmonary administration of HCQS suggests that pulmonary administration may produce higher tissue C_max an_d time above IC_50 wi_th overall low systemic exposure (59). To the best of our knowledge this is the first human aHCQS dose escalation study performed with a commercially available nebulizer following a traditional maximum tolerated dose study design, with ongoing safety board monitoring and PK assessments that are able to provide insight on distribution (60, 61). Since completion of our study, Hawari et al. (62) reported that aHCQS in humans was safe and associated with rapid absorption into the systemic circulation, both of which are consistent with our findings. The plasma levels they reported and the duration of drug exposure were, however, both much lower than we found, even with dose-adjustment, which probably reflects their use of a plasma assay, which is known to measure only a fraction of HCQ present in whole blood.

It is possible that aHCQS may provide benefit to disorders other than COVID-19, especially the many pulmonary inflammatory diseases for which current therapeutic options are limited. Importantly, our single-dose data in rats demonstrated that, compared to oral administration, cardiac HCQ concentrations were lower and lung parenchyma tissue concentrations were higher following IT administration, suggesting that aHCQS may have a greater margin of safety (Figure 2).

It is likely that there are multiple compartments within the respiratory tract cells into which HCQ distributes and that it takes sustained plasma or ELF levels to fill the slowly loading and emptying compartment(s) that account for the remarkably prolonged elimination of IV and oral HCQS. If that is the case, multiple doses of aHCQS may be required to attain high enough respiratory tract concentrations, along with prolonged ongoing exposure, to achieve an antiviral effect. In addition, the rapid absorption of aHCQS into the systemic circulation we observed may be driven by the large gradient between respiratory tract and blood concentrations. Narrowing that gradient by treating with oHCQS before aHCQS may result in higher and more sustained respiratory tract tissue concentrations and thus permit the loading of the potentially slow releasing compartment(s). Combining oHCQS and aHCQS would however increase the potential for cardiac toxicity.

Our studies have a number of limitations. 1. Prediction of tissue concentrations after administration of aHCQS to humans is complex and speculative because there are no available direct measurements of human HCQ tissue:blood ratios as a function of time. Although plasma concentrations of HCQ may be a better indicator of the potential efficacy and safety of aHCQS, our PK analyses were based exclusively on the whole blood concentrations in the human and rat studies because at some timepoints in the human Phase 1 study, HCQ was quantifiable in the whole blood, but not the plasma sample, and at low blood concentrations of HCQ there was considerable variability in the whole blood-to-plasma ratios (Supplementary Figure 2), probably reflecting variable binding to blood cells and platelets, as well as technical variability. Similarly, variability was also observed in the whole blood-to-plasma ratios in the rat studies (Supplementary Figure 2), where technical aspects relating to the blood drawing may have contributed to the variability. These observations are in line with published data on the difficulties of analyzing HCQ plasma levels (7). 2. Since the only data on the ratios of tissue:blood HCQ concentrations derive from the studies that we performed in rats (Table 1), we used them to estimate the tissue concentrations in humans. The applicability of this approach, however, assumes the following: i. Lack of between-species differences in dose deposition; ii. Lack of between-species differences in dose delivery; iii. Tissue concentrations, which were reported in µg HCQ/g tissue, can be converted to µg/mL.

Lethal COVID-19 is comprised of an early phase in which viral replication occurs in the lung, followed by an inflammatory/thrombotic phase that compromises oxygenation (56, 63–65). Thus, achieving drug concentrations with antiviral activity in the airways early in the course of disease is essential for improving clinical outcome, a premise supported by the need for early therapy with both monoclonal antibodies and the antiviral agents (66). Our first human-use experience with aHCQS provides data that it may be possible to rapidly produce high drug levels in respiratory tissue using a well-tolerated aerosol delivery method and thus may be applicable to other agents currently under development.

Finally, our first-in-human aHCQS dose escalation study, performed with a commercially available nebulizer, provides important tolerance and PK data on aHCQS that may inform the use of aHCQS in other pulmonary disorders in which its anti-inflammatory and immunomodulatory effects may be beneficial. Since HCQ manifests a number of complex PK features, and since HCQ metabolites may contribute to its observed beneficial and toxic effects, PK modeling without accounting for the metabolites and their biological activities may not be a reliable guide to likely clinical utility. As a result, carefully designed clinical trials are essential.

## Methods

### Animal Studies

A single dose study of the PK and tissue distribution of HCQ following oral (PO), intravenous (IV), and intratracheal (IT) administration of HCQS in male Sprague Dawley rats was performed. IT doses were administered as a solution instilled into the trachea.

All HCQ doses were adjusted using standard intraspecies scaling methods to be equivalent to standard human doses for each route of administration (67). As a prelude to the human studies, we characterized the systemic HCQ exposure by measuring plasma and whole blood concentrations and tissue distribution in the lung (large airway and parenchyma) and heart (left heart, right heart). One objective was to evaluate lung and heart exposure to HCQ when delivered at human equivalent doses via these different routes of administration.

A multiple dose study of the impact of intratracheal (IT) and intranasal (IN) HCQS solution on the Golden Syrian hamster preclinical model of SARS-CoV-2 infection was also performed. Syrian hamsters are susceptible to SARS-CoV-2 infection, developing reversible weight loss and interstitial pneumonia correlating with infectious dose (36). We studied the impact of IT and IN HCQS solution at high and low doses on this model (See Supplementary Methods).

The aHCQS investigational new drug enabling toxicology studies were performed in rats and beagle dogs, in compliance with Good Laboratory Practices (15).

### Human Phase 1 study

#### Objectives and Outcomes

The study’s primary objective was to assess the safety and tolerability of aHCQS administered as a single dose by oral inhalation with nasal exhalation (to enhance deposition in multiple airway regions) to healthy volunteers at escalating doses, starting with 20 mg, until either the maximum tolerated dose was identified, or 50 mg was administered. The secondary objective was to characterize the PK of a single dose of aHCQS in healthy adult volunteers. The study was approved by the Rockefeller University Institutional Review Board.

#### Study Population

10 healthy volunteers were enrolled after obtaining informed consent. To be eligible to participate, candidates had to be older than 18 years, non-smokers, and free of any chronic or acute respiratory illness. A complete list of inclusion and exclusion criteria is provided in the Supplementary Materials.

#### Study Design

This was a randomized, double-blind, placebo-controlled, single-dose, dose-escalation study in which 2 sentinel participants were first administered aHCQS in a single-blind manner in each cohort, and then, following assessment by the safety review committee, the dose was either escalated or 6 additional participants were randomized 4:2 to aHCQS versus placebo, respectively, in a double-blind manner. This trial was registered with ClinicalTrials.gov (NCT 04461353).

#### Randomization and Masking

After eligibility was confirmed and written informed consent obtained, participants were assigned either to a sentinel dosing group (single-blind) or according to double-blind randomization (aHCQS 4: Placebo 2). The study investigators, all research teams, and study participants were masked to treatment allocation, except for the sentinel dosing groups, in which case only the study participants were blinded. aHCQS and placebo were identical in appearance. The study medications were presented as ready-to-use aqueous solutions in syringes labeled according to regulatory requirements. A Cepacol Extra Strength Sore Throat Lozenge was administered 10 minutes before the inhalation to mask the bitter taste of aHCQS, and thus support blinding, and prevent cough.

#### Study Drug Dose and Administration via Nebulizer

Active doses were prepared by mixing the appropriate volume of sterile isotonic 100 mg/mL aHCQS solution with sterile 0.9% sodium chloride solution to achieve a total volume of 1 mL. Placebo doses were prepared using 1 mL of sterile Sodium Chloride Inhalation Solution USP 0.9% (Nephron Pharmaceuticals). Active and placebo doses of aHCQS (cohort 1: 1 mL of 20 mg/mL solution, cohort 2: 1 mL of 50 mg/mL solution) were prepared by an unblinded pharmacist, with all procedures and materials double verified. The study drug was administered by inhalation through the mouth using the Aerogen Solo\Ultra vibrating mesh nebulizer inhalation system (see Supplementary Figure 1 and Supplementary Materials for rationale of using the Aerogen nebulizer) and exhalation through the nose. All volunteers were shown a video explaining how to use the inhalation system. The volunteers were then instructed to inhale the study drug through the device’s mouthpiece and exhale through the nose until completion of the inhalation when no aerosol generation was observed. The volunteers were told that the duration of inhalation is usually less than 15 minutes and that they would be allowed to take pauses if needed.

Procedures to mitigate potential risk of infection with SARS-CoV-2 during the inhalation or while completing other study assessments that may be aerosol generating are provided in the Supplementary Methods.

#### Dose Selection and Breath Simulator Studies

The starting dose of aHCQS and the dose escalation plan for the study were based on the preclinical animal studies and the safety data from the previous clinical development program (15, 17). To verify the published data on the lung deposition achieved with the Aerogen inhalation system (68), and to obtain estimates of the inhaled dose of aHCQS (i.e., the dose that is nebulized and inhaled) and the regional distribution of the dose, we studied the delivery of aHCQS with the nebulizer connected to a breath simulator (see Supplementary Materials) (69).

#### Study Assessments

Volunteers were kept under observation for 6 hours after study drug administration and came back for assessments at 24 (±6) hours and 7 (±1) days after drug administration. Assessments included clinical evaluations, complete blood counts, serum chemistry, including liver function tests, urinalysis, pulmonary function tests (PFTs) [forced vital capacity (FVC) and FEV_1],_12-lead ECG, questionnaires related to sensory measures (smell and taste), drug and inhalation device acceptability, and cough assessment. The sensory measures were evaluated using a sensory measures, acceptability, and cough assessment questionnaire completed within 3 hours after inhalation, and a cough assessment questionnaire completed on days 2 and 8. Blood PK assessments of participants receiving aHCQS were made at baseline (before administration of the study drug) and at specified timepoints [+2, +3, +5, and +15 minutes, +1, + 2, +4, +6, and +24 (±4) hours, and +7 (±1) days].

#### PK Assessments and Analysis

HCQ levels were assayed by liquid chromatography-mass spectrometry/mass spectrometry (LC-MS/MS) on samples of whole blood anticoagulated with EDTA and on samples of plasma that were prepared from whole blood anticoagulated with EDTA by centrifugation at 1,500 x g for 10 minutes at 4°C. Samples were frozen at -80°C until analysis (whole blood at Altasciences and plasma at NorthEast BioLab). PK parameters were estimated by noncompartmental analysis using Phoenix WinNonlin software. Estimations of HCQ concentrations achieved immediately after inhalation in the epithelial lining fluid (ELF) were calculated using Mimetikos Preludium, ver. 1.1.7.1 based on published aerosol deposition patterns (70). The estimated respiratory tissue exposure was calculated by multiplying the HCQ concentration in humans by the observed tissue:blood HCQ concentration ratios at different timepoints in rats.

### Animal Studies

See Supplementary Methods.

#### Statistical Analysis

All preplanned summaries were descriptive in nature and no statistical comparisons were planned for primary and secondary analyses. Additional exploratory analyses that were not prespecified included comparisons between groups with the *t*-tests or one-way ANOVA including accounting for repeated measures and correction for multiple comparisons with Dunnett’s method, using a p-value of 0.05 to declare statistical significance. All exploratory statistical analyses were conducted with GraphPad Prism version 8.4.3.

#### Study approval

The Phase 1 study was approved by the Rockefeller University Institutional Review Board. Written informed consent was received prior to participation. All animal studies were approved by the local ethical committees.

## Data Availability

All data produced in the present study are available upon reasonable request to the authors.

## Data availability

Requests for data may be sent to the corresponding author.

## Author contributions

B.L.C., R.B.M., I.G. and B.S.C. conceived the project. O.S.B. designed and conducted the human studies, analyzed the data from the animal and human studies, and wrote the manuscript. B.S.C. and R.B.M critically revised the manuscript. R.H., D.B., and A.C.K. participated in planning and carrying out of the Phase 1 study. I.G. and R.B.M. and B.L.C. designed the preclinical studies and assisted in analysis of the results and developed the dosing regimens in humans. P.B. performed the drug deposition and physiologically based biopharmaceutical modeling. All authors participated in the design of the Phase 1 study, and critically reviewed and approved the manuscript.

## Acknowledgments

The authors thank the study participants and research teams at the Center for Clinical and Translational Science at the Rockefeller University and the Rockefeller University Hospital, as well as Robert S. Hillman, Oana Cociorva, Lisa G. Berdan, and MariJo Gleeson at Pulmoquine Therapeutics, Inc.

## ABBREVIATIONS

ACE2: Angiotensin converting enzyme 2
aHCQS: Aerosolized hydroxychloroquine sulfate
AUC: Area under the curve
Cmax: Maximum concentration
ECG: 12-Lead electrocardiogram
ELF: Epithelial lining fluid
FVC: Forced vital capacity
cGAS: Cyclic guanosine adenosine monophosphate
EUA: Emergency Use Authorization
FDA: Food and Drug Administration
HCQ: Hydroxychloroquine base
HCQS: Hydroxychloroquine sulfate
HR: Hour
IFN: Interferon
IN: Intranasal
IT: Intratracheal
IV: Intravenous
LC-MS/MS: liquid chromatography-mass spectrometry/mass spectrometry
Min: Minutes
oHCQS: Orally administered hydroxychloroquine sulfate
PK: Pharmacokinetic
PFT: Pulmonary function testing
PO: Oral
QTc: QT segment (corrected)
ROA: Route of administration
S: Spike
SAE: Serious adverse events
SRC: Safety Review Committee
TLR: Toll-like receptor
Tmax: Time of maximum concentration
TMPRSS2: Transmembrane serine protease 2
WHO: World Health Organization

