## Supplementary Material for "Preclinical and Human Phase 1 Studies of Aerosolized Hydroxychloroquine: Implications for Antiviral COVID-19 Therapy"

**Table of Contents**

Introduction

Rationale for developing aerosolized hydroxychloroquine sulfate (aHCQS) for delivery with the Aerogen Inhalation System for prevention and treatment of COVID-19

Methods

Animal Studies:

Pharmacokinetic and tissue distribution profile of hydroxychloroquine (HCQ) after single oral, intravenous, and intratracheal administration of HCQ Sulfate (HCQS) solution in male Sprague Dawley rats

Impact of intratracheal and intranasal HCQS solution on the Golden Syrian hamster preclinical model of SARS-CoV-2 infection

Human Studies:

Selection of Study Population: Inclusion and Exclusion Criteria

Procedures to mitigate potential risk of infection with SARS-CoV-2

Sensory measures, acceptability, and cough assessment questionnaires

Schedule of Assessments

In Vitro Evaluation of the Inhalation Delivery of aHCQ (Breath Simulator)

Results

In Vitro Evaluation of the Inhalation Delivery of aHCQ (Breath Simulator)

Supplementary Tables and Figures

### Introduction

### Rationale for developing aerosolized hydroxychloroquine sulfate (aHCQS) for delivery with the Aerogen Inhalation System for prevention and treatment of COVID-19

#### Rationale for developing aHCQS for prevention and treatment of COVID-19

Key reproducible observations on hydroxychloroquine sulfate (HCQS) and its potential role in preventing and treating COVID-19 include: i. Hydroxychloroquine (HCQ) is active against SARS-CoV-2 in vitro (Supplementary Table 1) (1-9); several publications reported that HCQ did not inhibit SARS-CoV-2 under certain experimental conditions (2, 6, 7, 9), but the maximal HCQ concentrations that were tested in those experiments were ≤100 µM; ii. When given in traditional systemic doses of ~200-400 mg per day, orally administered HCQS (oHCQS) is a safe and well-tolerated drug (10). The mechanism of action of HCQ against SARS-CoV-2 has not been fully elucidated. An increase in endosomal pH in the target cell is believed to adversely impact multiple stages of viral entry, replication, and budding (e.g., membrane fusion, acid-dependent proteolytic cleavage of the virus' S protein, post-translational modification of envelope glycoproteins), in addition to inhibition of glycosylation of the cell surface viral receptor, angiotensin-converting enzyme 2 (ACE2). It was also hypothesized that HCQ's anti-inflammatory effects play a role in mitigating the acute respiratory distress syndrome and cytokine storm that are associated with COVID-19 (11-14).

Clinical trial data demonstrated that none of the pharmacokinetic (PK) models that predicted that oHCQS would be effective in COVID-19 (4, 15, 16) translated into clinical efficacy, and this may be attributed to the following (15-18): i. The half-maximal inhibitory concentration ($IC_{50}$) values that were entered into PK models were observed in African green monkey kidney epithelium Vero E6 and Huh-7 cells (0.72-17.3 μM) (1, 3-6, 8, 9), whereas the $IC_{50}$ in human lung carcinoma-derived Calu-3 cells was substantially higher and estimated at 119 μM (Supplementary Table 1) (2); ii. A high lung to plasma coefficient (Kp = 541) was assumed from the first day of treatment with oHCQS in the first published PK model (4), even though Kp is only achieved at steady-state equilibrium, which requires several months of treatment with oHCQS (17, 19, 20), and even then, the only Kp values that are available in the literature are based on data from animal (19), rather than human, studies; iii. Estimated lung intracellular HCQ concentrations, which were compared to the in vitro $IC_{50}$ values, were the product of free unbound plasma concentration and Kp, but considering HCQ's sequestration in intracellular organelles and protein binding, the concentration actually available for antiviral activity is more likely only  the unbound plasma concentration (18, 21); iv. Scaling of in vitro $IC_{50}$ values (i.e., the drug concentration added to the culture supernatant) to whole blood and plasma concentrations is of uncertain relevance because the degree of drug binding to the cells in culture is unknown and both the intra-individual and inter-individual variability of the ratio of whole blood-to-plasma concentration of HCQ is high (15, 22, 23).

While most tInhaled drug therapy is used for targeting the lung while minimizing systemic exposure in order to improve the benefit/risk ratio of treatments for respiratory disease (27). Targeted delivery of HCQS or other antiviral therapies to the respiratory tissue (airways and lungs) via inhalation may hasten the onset of the effect and thus help achieve these goals in COVID-19 (28-35) and other respiratory infections and/or inflammatory diseases. Due to the relatively small volume of distribution of the lung compared to the rest of the body, the amounts of HCQS required to achieve local therapeutic concentrations are much less than those needed via systemic administration. Adding nasal exhalation to oral inhalation may also be important since the nose is usually the first human organ invaded by the SARS-Cov-2 virus, the highest expression of ACE2, the receptor that mediates SARS-CoV-2 entry into cells, is in the nose (36, 37) and high levels of active SARS-CoV-2 replication and viral shedding occur in the upper respiratory tract tissues during the first weeks of symptoms (24, 25).

Based on animal studies of other inhaled medications (38), inhalation of HCQS may result in achieving maximum concentrations in lung epithelial cells nearly instantaneously, with concentrations 100-fold or higher than those achieved after systemic administration. A previous clinical development program of aerosolized HCQS (aHCQS) for the treatment of asthma demonstrated that the treatment had a favorable safety profile in both healthy volunteers and patients with asthma, even when administered daily for 21 days (28, 29). These considerations led us to develop a novel formulation of aHCQS and study its safety, tolerability, and pharmacokinetics in humans.

Previous studies with aHCQS and rationale for selection of the Aerogen Inhalation System

A previous clinical development program of aHCQS for the treatment of asthma demonstrated a favorable safety profile in both healthy volunteers and patients with asthma, even when administered daily for 21 days (28, 29). That program was halted because the treatment did not demonstrate efficacy in treating asthma. The inhalation system used in those studies, AERx, required considerable training and cannot be connected to supplemental oxygen or a ventilator (39), so it would not be suitable for use by acutely ill respiratory patients or by patients who require supplemental oxygen or mechanical ventilation. Moreover, the AERx inhalation system was designed to deliver drug primarily to the alveoli rather than the large airways and nasal lining, whereas drug delivery to the latter regions is preferred to treat COVID-19 (24, 25, 36, 37). As a result, we chose the Aerogen inhalation system because it produces aerosols with a median volume droplet diameter of 4.3 – 5.2 microns, suitable for upper and central airway deposition (40). In addition, the system can be used in both the inpatient and outpatient settings, including patients who are already on a ventilator.

### Methods

#### Animal Studies

Pharmacokinetic and tissue distribution profile of HCQ after single oral (PO), intravenous (IV), and intratracheal (IT) administration of HCQS solution in male Sprague Dawley rats

These experimental procedures were performed at Truly labs (Sweden) and were approved by the local ethical committee before initiating the study. The aim of these experiments was to evaluate the PK and blood, heart, and respiratory tissue distribution of HCQ after systemic PO, IV, and IT dosing of HCQS solution to rats. HCQS solution in 0.9% sodium chloride was administered by oral gavage (PO), IV injection, or IT instillation to male Sprague Dawley rats (age 6 weeks, weight 290-372 g). Each treatment group included 6-8 rats. The study was designed to approximate weight-normalized human doses used in trials of orally administered HCQS (oHCQS) for COVID-19. Doses were scaled to species as per Tepper et al. (42). For the PO dose, the human oHCQS loading dose of 800 mg was scaled to an adult human body weight of 60 kg (13.3 mg oHCQS/kg). For IV injection, an approximately equivalent systemic dose of 9.9 mg/kg was selected assuming oral bioavailability to be ~74% (22). For IT administration, the dose was calculated based on a human lung dose of HCQS of 30 mg (0.03 mg/g lung tissue for the human lung weight, 1,000 g; 0.18 mg/kg body weight for a rat weighing 0.25 kg with a lung weight of 1.5 g). The concentrations of the formulation were adjusted so that they translated into volumes of administration customary for rat experiments (Supplementary Table 2). Before dosing, the rats were weighed and marked for identification. The rats were conscious during IV and PO administration. IT administration was performed under isoflurane anesthesia. When the rat reached sufficient anesthetic depth, it was placed in a supine position on a board tilted approximately 45 degrees with its head up. The IT administration was done with a syringe connected to a blunt steel cannula (with a small marble at the top to prevent injury to the airways). The cannula passed the larynx, and the administration was made when the rings of cartilage in the trachea could be felt. The rats were allowed to wake up in the supine position. The syringes were weighed before and after dosing to obtain the actual delivered dose (PO: 13.2-13.9 mg/kg, IV: 9.9-10.3 mg/kg, IT 0.15-0.24 mg/kg). Each rat was sampled for blood and plasma from 1 to 10 times depending on the time of termination. Blood was collected from the sublingual vein (200 µL/sample) into lithium-heparin pre-coated microvette tubes. Plasma (produced by centrifugation at 2,500 x g for 5 minutes at 4°C) and whole blood were promptly frozen at -80°C. At termination and after the last blood sample was collected, the rats were anaesthetized with isoflurane and the lungs were perfused with 0.9% sodium chloride using a catheter inserted in the right ventricle until all visible blood was cleared from the lungs. The heart was then removed and cut in two halves (left and right atrium/ventricle), weighed, and put on dry ice. The lung parenchyma was removed from the large airways using a blunt spatula. The parenchyma from one of the right lung lobes was weighed and put on dry ice. After removal of the trachea at the level of the bifurcation, the large airways were weighed and put on dry ice. Tissue samples were weighed and then homogenized (2.5 mL of acetonitrile was added to the sample in GentleMACS M tubes, and the samples were homogenized using a GentleMACS Dissociator [Miltenyi Biotec]). The following samples were sent for bioanalysis of HCQ levels by liquid chromatography-mass spectrometry/mass spectrometry (Recipharm, Sweden; lower limit of detection 1 ng/mL HCQ): whole blood, plasma, left heart, right heart, parenchyma from one of the right lobes, and large airways with trachea removed at the bifurcation. PK parameters were estimated where possible by noncompartmental analysis using Phoenix WinNonlin software.

Impact of Intratracheal (IT) and intranasal (IN) HCQS solution on the Golden Syrian hamster preclinical model of SARS-CoV-2 infection

These experimental procedures were performed at Bioqual, Inc. (Rockville, MD) and were approved by the local ethical committee before initiating the study. Syrian hamsters are susceptible to SARS-CoV-2 infection, developing reversible weight loss and interstitial pneumonia correlating with infectious dose (8, 43, 44).

The aim of these experiments was to study the impact of combined IT and intranasal (IN) HCQS solution on weight loss, SARS-CoV-2 viral load in oral swabs on days 2, 4, and 7 post-infection, and lung histopathology in Syrian hamsters (age 6-8 weeks, weight 82-113 g) infected with SARS-CoV-2 via IN inoculation (See Supplementary Table 3). The same formulation of HCQS that was used in the rat study was used for these studies, but the dose was calculated based on a human dose of HCQS of 60-120 mg loaded in the nebulizer using the allometric scaling conventions for rats since there are no established conventions for hamsters (42). The HCQS dose was diluted with 0.9% sodium chloride to a total volume of 10 μL for IT dosing and 100 μL for IN dosing. The study design is summarized in Supplementary Table 3.

### Human Studies

#### Selection of Study Population

##### Inclusion Criteria

Each study participant had to meet all of the inclusion criteria listed below to be eligible to participate in this study.

1. Willing and able to give written informed consent.
2. Males or females aged ≥18 years old.
3. Good general health as determined by no acute illness and no clinically significant abnormal findings on medical history, vital signs, laboratory tests, or physical examination at screening that, in the opinion of the PI, would interfere with study drug administration, jeopardize the safety of the study participant, or impact the validity of the study results; participants with stable chronic illness are allowed at the discretion of the PI.

4. An interpretable 12-lead ECG with a corrected QT (QTc) interval ≤450 ms, according to Bazett's formula, without evidence of clinically significant abnormal findings.

5. Normal $FEV_1/FVC$ ratio, defined as any value above 0.7 or above the lower 5th percentile of normal AND $FEV_1$ >80% of predicted or above the lower 5th percentile of normal.

6. Pulse oximetry $0_2$ saturation ≥95% in room air.

7. Negative test result for COVID-19 within 7 days of Day 1 AND concurrent with local hospital policy:

   - A nasopharyngeal swab tested with the ID NOW COVID-19 assay (Abbot). OR

   - A negative RNA-based test result of an oropharyngeal or nasopharyngeal swab or saliva sample performed according to CLIA/CLEP.

8. Females of child-bearing potential must be non-pregnant, non-lactating, have a negative urine pregnancy test at screening, and agree to use an acceptable form of birth control for 200 days after the last administration of the study drug. Females are considered of non-childbearing potential if they are postmenopausal (last menstrual period at least 1 year before screening) or have been surgically sterilized (documented hysterectomy, tubal ligation, or bilateral oophorectomy) for at least 6 months at screening.

9. Willing to comply with protocol-defined procedures and complete all study visits.

10. Willing to use the Inhalation System and exhale through the nose.

11. Adequate venous access in the left or right arm to allow collection of required blood samples.
12. Participant understands and communicates in English.
13. Serum Potassium level ≥3.5 mEq/L, Serum Magnesium level ≥1.5 mg/dL, and Serum Calcium ≥8.5 mg/dL.

Exclusion Criteria

Each study participant could NOT meet any of the exclusion criteria listed below to be eligible to participate in this study.

1. Any self-reported symptoms of influenza-like or COVID-19-like illness in the 14 days preceding the study visit: Fever >101.4 °F, sore throat, nasal congestion, post-nasal discharge, shortness of breath, gastrointestinal distress, wheezing, cough, headache, or fatigue.
2. Any history of diagnosed chronic lung disease, including but not limited to asthma or chronic obstructive lung disease.
3. Symptoms of seasonal allergies or use of any drugs for seasonal allergies or any inhaled (oral/nasal) drugs in the 2 weeks prior to Day 1. Mild seasonal allergy symptoms that have not altered sleep or activity patterns nor required use of over-the-counter (OTC) or prescription medications are allowed.
4. Any close contact exposure in the past 28 days to a person who was diagnosed as having COVID-19, with or without laboratory confirmation, during that close contact exposure or in the ensuing 14 days OR a similar encounter with a person who was determined to have suspected COVID-19, defined by that person being

ordered to enter isolation for that indication by a medical authority. Close contact is defined as being within approximately 6 feet of a COVID-19 case for a prolonged (>10 minutes) period of time and can occur while caring for, living with, visiting, or sharing a healthcare waiting area or room with a COVID-19 patient OR having direct contact with infectious secretions of a COVID-19 patient (e.g., being coughed on), if such contact occurred while not wearing the recommended personal protective equipment for that type of contact [e.g., gowns, gloves, N95 respirator (or equivalent), eye protection].

5. Any participant with a history of SARS-CoV-2 infection that was confirmed by testing or diagnosed without testing within 4 weeks preceding Day 1. If infection occurred more than 4 weeks prior, candidates may be enrolled if they meet the rest of the eligibility criteria.

6. Any participant with a history severe respiratory illness that required hospitalization in the 60 days preceding Day 1 OR any participant with a history severe respiratory illness that required hospitalization in the preceding 120 days without full recovery.

7. Participation in another clinical study that involved treatment with an investigational product or device within 30 days of screening or during the study.

8. Participants with a known history of human immunodeficiency virus (HIV) infection.

9. Known, active hepatitis A, B, or C infection.

10. History of bronchospasm in response to use of an inhalation device.

11. Use of any prescription medication (except oral contraceptives) during the 30 days prior to study dosing that may affect drug absorption, metabolism and excretion, prolong the QTc interval, affect drug efficacy, or increase the risk of adverse reactions, unless approved by the Principal Investigator.
12. Use of any OTC product, herbal product, diet aid, hormone supplement, etc., within 7 days prior to dosing unless approved by the Principal Investigator.
13. Unwilling or unable to provide written informed consent.
14. Any known hypersensitivity to quinolines (e.g., hydroxychloroquine, chloroquine, primaquine, quinine) or known history of glucose-6-phosphate dehydrogenase (G6PD) deficiency or any contraindication to oral hydroxychloroquine.
15. Known retinopathy, fundus disease, or macular disease.
16. Diagnosis of long QT Syndrome.
17. Smoking of tobacco or non-tobacco substances, or vaping, within the last 6 months.
18. Severe obesity (body mass index [BMI] ≥35 kg/$m^2$).

PI = Principal investigator; ECG = Electrocardiogram; $FEV_1$ = Forced Expiratory Volume in 1 second; FVC = Forced Vital Capacity; CLIA/CLEP = Clinical Laboratory Improvement Amendments / New York Clinical Laboratory Evaluation Program

<u>Procedures to mitigate potential risk of infection with SARS-CoV-2</u>

In order to mitigate any potential risk of infection with SARS-CoV-2, considering that there were concerns of false negative testing for SARS-CoV-2 early in the pandemic (45,

46), and since inhalations and pulmonary function tests (PFTs) may be considered aerosol generating procedure (AGPs) (47-51), these procedures were performed in an airborne infection isolation room and staff donned personal protective equipment appropriate for risk of an airborne infection. Although the Aerogen inhalation system has a relatively low concentration of fugitively-emitted aerosols (52) a filter (AirLife, CareFusion) was employed on the exhalation port of the nebulizer mouthpiece for extra environmental safety. In addition, study volunteers were required to test negative for active SARS-CoV-2 infection within 24 hours of study visits.

Sensory Measures, Acceptability, and Cough Assessment Questionnaire (Day 1)

The following questionnaire was filled out by participants within 3 hours after completion of study drug administration on Day 1:

**SMELL**

**Please rate the smell by ticking ONE box.**

| 0 | 1 | 2 | 3 | 4 | 5 |
|---|---|---|---|---|---|
| ☐ | ☐ | ☐ | ☐ | ☐ | ☐ |
| No smell | Slight smell, not unpleasant | Slight smell, unpleasant | Moderate smell, unpleasant but tolerable | Strong smell, unpleasant but tolerable | Very strong smell, intolerable |

**TASTE**

**Please rate the taste by ticking ONE box.**

| 0 | 1 | 2 | 3 | 4 | 5 |
|---|---|---|---|---|---|
| ☐ | ☐ | ☐ | ☐ | ☐ | ☐ |
| No taste | Slight taste, not unpleasant | Slight taste, unpleasant | Moderate taste, unpleasant but tolerable | Strong taste, unpleasant but tolerable | Very strong taste, intolerable |

**BITTERNESS**

**Please rate the bitterness by ticking ONE box.**

| 0 | 1 | 2 | 3 | 4 | 5 |
|---|---|---|---|---|---|
| ☐ | ☐ | ☐ | ☐ | ☐ | ☐ |
| Not bitter | Slightly bitter, not unpleasant | Slightly bitter, unpleasant | Moderately bitter, unpleasant but tolerable | Strongly bitter, unpleasant but tolerable | Very strongly bitter, intolerable |

Please answer these questions regarding the inhalation of the study drug:

1. Please rate the taste of the study medication:

☐ Very unpleasant

☐ Unpleasant

☐ Somewhat unpleasant

☐ Neutral

☐ Somewhat pleasant

☐ Pleasant

☐ Very pleasant

Comments: ___________________________________________

2. I would be willing to use this study medication on a regular basis.

☐ Strongly Disagree

☐ Disagree

☐ Somewhat Disagree

☐ Neutral

☐ Somewhat Agree

☐ Agree

☐ Strongly Agree

Comments: ___________________________________________

3. The taste of the study medication would not keep me from using it on a regular basis.

☐ Strongly Disagree

☐ Disagree

☐ Somewhat Disagree

☐ Neutral

☐ Somewhat Agree

□ Agree

□ Strongly Agree

Comments: ___________________________________________

4. Overall, I would not have a problem using the study medication on a regular basis.

□ Strongly Disagree

□ Disagree

□ Somewhat Disagree

□ Neutral

□ Somewhat Agree

□ Agree

□ Strongly Agree

5. Please rate how easy it was for you to use the Aerogen device:

□ Very Difficult

□ Difficult

□ Somewhat Difficult

□ Neutral

□ Somewhat Easy

□ Easy

□ Very Easy

6. Please mark a red dot just above this line to characterize any cough that you may have had starting with the inhalation of the study drug until the completion of this questionnaire:

No cough

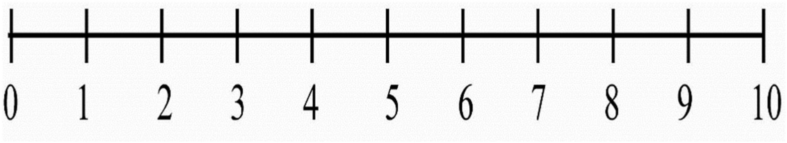

Worst cough ever

7. What was the total duration of cough that you had since starting the inhalation of the study drug?

☐ All of the time

☐ Most of the time

☐ A good bit of the time

☐ Some of the time

☐ A little of the time

☐ Hardly any of the time

☐ None of the time

8. During the inhalation of the study drug, were you able to exhale through the nose as you were encouraged:

☐ All of the time

☐ Most of the time

☐ A good bit of the time

☐ Some of the time

- ☐ A little of the time
- ☐ Hardly any of the time
- ☐ None of the time

Cough Assessment Questionnaire (Day 2, Day 8)

The following questionnaire was filled out by participants on Day 2 (24±6 hours after administration of the study drug) and Day 8 (7±1 days after administration of the study drug):

1. Please mark a red dot just above this line to characterize any cough that you may have had since you completed the Day 1 questionnaire:

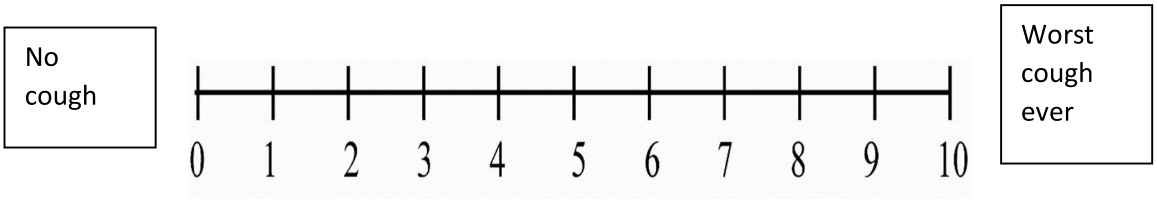

2. What was the total duration of cough that you had since you completed the Day 1 questionnaire?

☐ All of the time

☐ Most of the time

☐ A good bit of the time

☐ Some of the time

☐ A little of the time

☐ Hardly any of the time

☐ None of the time

Schedule of Assessments

| Study Procedures | Day -7 to -1[1] (Screening) | Day 1 (Treatment) | Day 2 | Day 8 (±1) | Day 30 (+7) |
|---|---|---|---|---|---|
| Evaluation of COVID-19 symptoms | X | X | X | X | X |
| Day Visit | | X | | | |
| Outpatient Visit | X | | X | X | |
| Phone call | | | | | X |
| Informed Consent | X | | | | |
| Collection of oropharyngeal or nasopharyngeal swab or saliva for COVID-19 RNA-based testing[2] | --------------------------X------------------------- |  |  |  |  |
| Inclusion/Exclusion | X | | | | |
| Demographics | X | | | | |
| Prior/Concomitant Medications[3] | X | X | X | X | X |
| Medical History | X | | | | |
| Physical Examination[4] | X | X | X | X | |
| Vital Signs[5] | X | X | X | X | |
| Height, weight, waist circumference, BMI[6] | X | X | X | X | |
| PFTs (FEV$_1$ and FVC)[7] | X | X | X | X | |
| 12-lead ECG[8] | X | X | X | X | |
| Randomization, if applicable[9] | | X | | | |
| Clinical Laboratory Tests | | | | | |
| Complete blood count (CBC) with Differential[10] | X | | | X | |
| Chemistry[11] | X | | | X | |
| Testing for (HIV, HBV and HCV) | X | | | | |
| Glucose by Glucometer (1-hour post-dose) | | X | | | |
| Urinalysis[12] | X | | | X | |
| Urine Pregnancy Test, if applicable | X | | | | |
| Prophylaxis with Cepacol and/or Ipratropium, if applicable[13] | | X | | | |
| Study Drug (aHCQS or placebo) | | X | | | |
| PK Blood Samples[14] | | X | X | X | |

| | | | | |
|---|---|---|---|---|
| Adverse Events[15] | ----------------------------------------X--------------------------------------- |  |  |  |
| Sensory Measures, Acceptability, and Cough Assessment Questionnaire[16] | | X | | |
| Cough Assessment Questionnaire | | | X | X |

1. Screening will be done within 7 days of the planned study drug administration, and may (partly) be performed pre-dose on Day 1.
2. Testing for COVID-19 will be performed in 7 days preceding the following potentially aerosol generating procedures: Screening PFTs, Day 1 PFTs, and inhalation of the study drug. Testing will be repeated on Days 2 to 8 only if participants report suspected symptoms of COVID-19, including but not limited to those listed in the eligibility criteria. Collection of sample and testing prior to screening and Day 1 may be waived if the participant provides the results of a negative test result with an RNA-based assay that was performed according to CLIA/CLEP within 7 days prior to performing screening PFTs and the inhalation of the study drug on Day 1. If there is an indication for repeated testing after Day 1, the test may be waived if the participant provides negative results of an RNA-based test that was performed according to CLIA/CLEP and after the onset of symptoms suspected for COVID-19. COVID-19 testing policy will also be concurrent with local hospital policy.
3. Prior medications taken within 6 months of signing the informed consent form (ICF) will be recorded. Concomitant medications, including nonprescription and herbal medications, will be collected at every visit beginning after written informed consent is obtained.
4. The physical examination at screening will consist of general appearance, HEENT, neurological, heart/cardiovascular, lungs, abdomen, endocrine, extremities, musculoskeletal, lymphatic, and skin; it will not include a rectal, gynecological, or genitourinary examination. A directed physical examination assessing and documenting changes from the previous visit, including any new abnormalities, will be conducted at on Day 1 (pre-dose, within 3 hours of dose), and Days 2 and 8. If the brief physical examination shows no changes from the previous visit, this must be documented.
5. Vital signs include respiratory rate, temperature, seated blood pressure, pulse, and $O_2$ saturation measurement by pulse oximetry, and will be recorded at screening and on Day 1 (pre-dose, within 30 minutes of initiation of dose administration), before PFTs and blood drawing, and after the participant is allowed to rest seated for at least 5 minutes. Post-dose vital signs will be recorded on Day 1 at approximately +10, +30 minutes, and +1, +2, +3, and +6 hours after completion of administration of the entire inhalation volume or the session administered dose for that participant. If a participant's dose is split into several administrations, vital signs will also be measured within 10 minutes after the participant completes the first part of the inhalation administration and before the participant initiates the next inhalation. Vital signs will also be measured once on Day 2 (+24 ± 6 hours post dose) and Day 8 after the participant is allowed to rest seated for at least 5 minutes. The first blood pressure measurement on each study visit will consist of 2 consecutive measurements of blood pressure ≥10 minutes apart. If any of the Day 1 post-dose blood pressure measurements differ by ≥25% from the mean value of the baseline readings for that date or if the participant coughs during the measurement, the measurement at that time point will be repeated. All measurements will be recorded.
6. Height and waist circumference will be measured and BMI will be calculated at screening only.
7. PFTs ($FEV_1$ and FVC) will be performed at screening and on Day 1 at pre-dose (within 25 minutes of dose) and at approximately +15 minutes, and +1, +3 and +6 hours after completion of administration of the entire inhalation volume or the session administered dose, and on Day 2 and Day 8.

8. 12-lead ECG will be performed at screening and on Day 1 pre-dose (within 3 hours of dose) and approximately +2 and +6 hours, and on Days 2 and 8. The ECG will be performed after the participant is allowed to rest seated for at least 5 minutes, before transferring to the examination bed/table for the ECG. The pre-dose and Day 2 and Day 8 ECG will be performed before PFTs or blood drawing.
9. The first 2 participants in each dose cohort will receive aHCQS in a single-blind manner (i.e., only the participants will be blinded to treatment assignment) on Day 1. The subsequent 6 participants in each dose cohort will be randomized 4:2 to aHCQS or placebo on Day 1.
10. Complete blood count (hematocrit, hemoglobin, mean corpuscular volume, RBC count, platelet count, WBC count with differential [neutrophils, lymphocytes, monocytes, eosinophils, basophils, other cells] drawn at screening and on Day 8.
11. Chemistry panel (albumin, total protein, ALP, ALT, AST, direct and indirect bilirubin, GGT, BUN, creatinine, glucose, bicarbonate, calcium, chloride, magnesium, phosphate, potassium, sodium, and LDH) drawn at screening and on Day 8.
12. Urinalysis (pH, specific gravity, protein, glucose, ketones, urobilinogen, bilirubin, leukocyte esterase, squamous cells, epithelial cells, clarity, bacteria, blood) will be obtained at screening and Day 8.
13. Prophylaxis for bitter taste and cough will only be administered to specific participants or dose cohorts, as described in the protocol. The order of prophylaxis administration will be Cepacol followed by Ipratropium. The study drug will be administered 10 minutes after completion of the final prophylaxis regimen.
14. Blood samples for PK analysis will be obtained on Day 1 pre-dose (time 0) and approximately +2, +3, +5, and +15 minutes after completion of administration of the entire dose of the inhalation, and also approximately +1, +2, +4 and +6 hours post-dose completion. A single blood sample for PK will be drawn on Day 2 (+24±4 hours post dose) and Day 8 (± 1 day).
15. Adverse events will be collected and recorded at every visit, beginning after written informed consent is obtained.
16. The Day 1 Sensory Measures, Acceptability, and Cough Assessment Questionnaire will be filled out by the participant within 3 hours of completion of the inhalation administration.

aHCQS = Aerosolized hydroxychloroquine sulfate; CLIA/CLEP = Clinical Laboratory Improvement Amendments / New York Clinical Laboratory Evaluation Program; ECG = Electrocardiogram; $FEV_1$ = Forced Expiratory Volume in 1 second; FVC = Forced Vital Capacity; PFT = Pulmonary function testing; PK = Pharmacokinetics

**In Vitro Evaluation of the Inhalation Delivery of aHCQ (Breath Simulator)**

These experimental procedures were performed at the School of Pharmacy in the University of Sydney, Australia, by Drs. Hak-Kim Chan and Philip Chi Lip Kwok.

Aerosolized hydroxychloroquine sulfate (aHCQS) delivery was simulated by administering aHCQS with the Aerogen Solo\Ultra inhalation system connected to a breath simulator according to the United States Pharmacopeia (USP) methods (53). Three new Aerogen Solo\Ultra nebulizers were used to perform these experiments. The same Aerogen controller was used for all experiments.

A 0.55% w/v solution of sodium chloride in deionized water was made as the diluent. 10 g of HCQS USP powder (Lot 1910P031, Batch 033600-192021; Sci Pharmtech Inc, Taoyuan, Taiwan) per 100 mL of the final volume was weighed into a volumetric flask. HCQS was dissolved by adding the 0.55% w/v sodium chloride diluent to approximately 60% of the final volume. 24 mL of 0.1 N sodium hydroxide was then added to the volumetric flask per 100 mL of the final volume. Small amounts of white precipitates initially appeared upon adding the sodium hydroxide solution but they quickly dissolved within a second. The final volume was made up with the sodium chloride diluent to obtain 100 mg/mL of HCQS. The 20 and 50 mg/mL HCQS solutions were prepared by diluting the 100 mg/mL solution accordingly with 0.9% sodium chloride.

HCQS dose output from Aerogen Solo\Ultra

One SureGard filter was connected to the outlet of the Aerogen Ultra mouthpiece. Filters were also fitted to the exhaust end of the mouthpiece and the exhaust port at the bottom of the Aerogen Ultra (Supplementary Figure 1). There were thus one outlet filter and two exhaust filters. Silicone adaptors were used to connect the mouthpiece to the outlet and one of the exhaust filters (Supplementary Figure 1). The experiments were conducted under ambient conditions (18-25°C, 20-65% relative humidity). The procedure followed the USP method except that the aerosols were collected from the start to the end of nebulization instead of collecting them for the first minute using one output filter and then collecting the rest of the aerosol with another output filter. This was to avoid drug loss when changing the filters. The end of nebulization was determined by visual inspection when no solution remained in the nebulizer. HCQS solution was added into the reservoir of the Aerogen Solo by pipetting. The PWG-33 breathing simulator (Piston Medical, Budapest, Hungary) was connected to the output filter. The simulated breathing waveform was sinusoidal at 15 cycles/minute, with an inhalation-to-exhalation ratio of 1:1 and a tidal volume of 500 mL. The outlet and exhaust filters captured droplets exiting the nebulizers during the inhalation and exhalation phases in the breathing cycle, respectively. The nebulizer and breathing simulator were operated from the start to the end of nebulization, after which the setup was left to stand for 20 minutes before being removed and assayed. This was to allow the droplets in the Aerogen Ultra to settle by gravitational sedimentation and avoid potential aerosol loss if the setup was disassembled immediately. The runs were conducted in triplicate for each nebulizer and 3 nebulizers were tested with each dose (20 mg/mL, 1 mL; 50 mg/mL, 1 mL; 100 mg/mL, 1 mL). The openings of the two exhaust filters were sealed with Parafilm after adding in 10 mL of deionized water. The exhaust filters were then exhaustively rinsed by shaking for 5 minutes. The outlet filter was placed into a 600 mL glass beaker. Four hundred mL of deionized water, a glass weight, and magnetic stirrer were added into that beaker afterwards. The glass weight was to weigh down the filter to ensure its complete immersion in the water. The mixture was magnetically stirred for 5 minutes, followed by shaking for another 5 minutes. The liquid reservoir and outlet of the Aerogen Solo were exhaustively washed with 10 mL of deionized water and 6 minutes of shaking in total. The same was performed on the two silicone adaptors. The washings were collected into a 100 mL volumetric flask. The openings of the Aerogen Ultra were sealed with Parafilm after adding about 10 mL of deionized water. The whole chamber was exhaustively rinsed as a whole in the same manner as described for the Aerogen Solo. All washings were pooled into the same volumetric flask. The volume was made up to 100 mL with deionized water. All samples were assayed by HPLC. HCQS was quantified by the modified reverse phase-HPLC method from the USP. The assay was performed on an automated HPLC system (Shimadzu, Kyoto, Japan) with an Agilent Zorbax SB-C18 column (5 μm, 4.6X250 mm; Waters, Milford, MA). The mobile phase was composed of 10:10:80 v/v methanol, acetonitrile, and 0.6 mM sodium 1-pentanesulfonate aqueous solution with 0.2% v/v phosphoric acid. The mobile phase and all the other solvents used in the HPLC were filtered and degassed before use. The column was kept at 35°C in a CTO-20A column oven (Shimadzu, Kyoto, Japan). Each sample ran for 15 min at 1 mL/min. The injection detection wavelength and volume were 254 nm and 20 μL, respectively. Standard solutions (6.25-1,000 μg/mL) were freshly prepared by serially diluting a 100 mg/mL HCQS solution aliquot that had been filtered through a sterile Millex-GP hydrophilic polyethersulfone membrane syringe filter (0.22 μm pore size; Millipore, Burlington, MA, USA). The diluent for the standard solutions was deionized water and 50:50 v/v methanol:water, depending on the diluent used for the samples. The sample solutions were diluted 10- to 1,000-fold with 0.9% sodium chloride when required to obtain absorbances within the range of the calibration curves. Each experiment was repeated 3 times.

Droplet size distribution measured by laser diffraction

The nebulized droplets were sized by laser diffraction using Spraytec (Malvern Panalytical, Malvern, UK) with an inhalation cell and at an acquisition frequency of 2.5 kHz. The outlet of the Aerogen Ultra mouthpiece was positioned 1 cm from the laser measurement zone to minimize evaporation during measurement. A vacuum pump connected to the other end of the inhalation cell was used to remove the aerosols continuously at 65 L/min to: 1) prevent re-entrainment of droplets into the laser measurement zone; and 2) keep the laser signal transmission >70% to minimize multiple scattering. The Aerogen Ultra mouthpiece was not sealed to the inhalation cell so the airflow through the Aerogen Ultra was unknown. Signals from Detectors 1-10 were excluded to account for beam steering effects. The real and imaginary refractive indices for the droplets were taken to be the same as those for water, which were 1.33 and 0.00, respectively. The refractive index for air was 1.00. These values were deemed appropriate because all measurements showed low residual values (<0.5%). The droplets were sized when the signal transmission was <99.9%. The duration of nebulization was the time that aerosols were seen by eye to traverse continuously through the laser measurement zone. The raw data was processed to yield an averaged volumetric diameter distribution for the nebulization period in a given run. The runs were conducted in triplicate for each nebulizer and 3 nebulizers were tested with each dose (20 mg/mL, 1 mL; 50 mg/mL, 1mL; 100 mg/mL, 1 mL). The relative humidity cannot be controlled due to physical and resource limitations. In preliminary experiments (data not shown) extra laser diffraction experiments were conducted for the 50 mg/mL and 100 mg/mL doses and no clear dependence between droplet size and relative humidity (27-60%) was observed.

Cascade impaction

Aerosol performance was measured by the USP method using a Next Generation Impactor (NGI; USP Apparatus 5) without a pre-separator. The runs were conducted in triplicate for each nebulizer and 3 nebulizers were tested with each dose (20 mg/mL, 1 mL; 50 mg/mL, 1 mL; 100 mg/mL, 1 mL). The NGI and throat were chilled at 5°C for at least 90 minutes before the experiment. After chilling, a SureGard filter was connected to the NGI after the micro-orifice collector (MOC) to capture any drug that passed beyond the lowest impactor stage. The sealing of the apparatus was verified before each run by a vacuum leak test, after which the airflow rate was set to 15 L/min. A silicone adaptor was used to connect the mouthpiece to the USP induction port (throat). The experiments were conducted under ambient conditions (18-25°C, 20-65% relative humidity). HCQS solution was added into the reservoir of the Aerogen Solo by pipetting. No exhaust filters were required to be connected to the Aerogen Ultra because the airflow was suction-only. The nebulizer and vacuum pump were operated from the start to the end of nebulization. The end of nebulization was determined by visual inspection when no solution remained in the nebulizer. The setup was left to stand for 20 minutes before being removed and assayed. The co-solvent used for all NGI samples was 50:50 v/v methanol:water. The Aerogen Solo and Aerogen Ultra were exhaustively washed with this co-solvent, collected into a 100 mL volumetric flask, and made up to volume. The post-NGI filter was washed with 10 mL of the co-solvent, as for the dose output exhaust filter. The adaptor, throat and NGI impactor stages were washed with 4 mL of the co-solvent. The assay for the 100 mg/mL HCQS runs was conducted in the same manner, except that Stages 1-6 (14.1 – 1.36 μm) were washed with 20 mL instead of 4 mL of the co-solvent. The metered dose was the amount of HCQS added into the nebulizer. The emitted dose was the total amount of drug assayed from the adaptor to the post-NGI filter. The recovered dose was the total amount of HCQS assayed on all the parts in the experimental setup, i.e., from the nebulizer to the post-NGI filter. Fine particle doses (FPDs) under 1, 2, 3, and 5 μm were calculated, from which the corresponding fine particle fractions (FPFs) with respect to the total metered, emitted, and recovered doses were then derived.

### Results

#### In Vitro Evaluation of the Inhalation Delivery of aHCQ (Breath Simulator)

For additional data on these experiments please refer to Tai W et al. (40). The total amount of drug predicted to be inhaled over the entire course of nebulization (dose output) of aHCQS (1 mL of 20 mg/mL, 50 mg/mL, and 100 mg/mL) with the Aerogen Solo\Ultra based on simulation studies in which the nebulizer was connected to a breath simulator is shown in Supplementary Table 6. The predicted inhaled dose was calculated by measuring the mass of HCQS captured by the filter that was connected to the output end of the mouthpiece and was ~50% of the metered dose (~50% of the metered dose was retained in the Aerogen Solo\Ultra). The total amount of drug at the exhaust filters was measured and summed and found to be <2% of the metered dose. A trend of increasing nebulization duration as a function of drug concentration was observed, going from 231 ± 33 seconds (mean ± SD) for 1 mL of 20 mg/mL to 306 ± 11 seconds for 1 mL of 50 mg/mL ($p<0.001$ compared to the 20 mg dose), and 384 ± 22 seconds for 1 mL of 100 mg/mL ($p<0.001$ compared to the 20 mg dose), consistent with published observations that solute concentration affects the nebulization time (54).

The droplet size distribution measured by laser diffraction was stable over the entire period of nebulization, with a volumetric median diameter between 4 and 5 μm for all concentrations and volumes (Supplementary Table 7, Supplementary Figure 3A). The droplet size distribution was not affected by the HCQS concentration or nebulizer unit tested in the study. The nebulization duration in the laser diffraction experiments was generally longer than in the dose output and NGI experiments because the visual determination of the end of nebulization was different between the experiments (the aerosol could be easily observed visually in laser diffraction but could not be seen clearly in the dose output and NGI experiments).

Aerosol performance (cascade impaction) was measured by the USP method using an NGI. Only a small amount of HCQS was collected on the post-NGI filter (<1%) so the NGI captured practically all the emitted doses. The total recovered dose was also close to the metered dose, indicating satisfactory recoveries. The overall aerosol performance profiles of the HCQS solutions were similar, with minimal throat deposition (Supplementary Table 7, Supplementary Figure 3B).

The fraction of the metered dose at the output filter that was observed in the dose output experiments was 44.2-49.7%, with ~50% of the dose remaining in the Aerogen Ultra\Solo and <2% at the exhaust filters. The laser diffraction experiments measured 47.9-58.6% of the aerosol at <5μm, i.e., respirable fraction. In the cascade impaction experiments the emitted dose was 66.0-67.6% of the metered dose and the fine particle (<5 µm) fraction of the emitted dose was 67.3-71.8%. The estimated lung dose based on these observations is between 21.2% (44.2% X 47.9%) and 48.5% (67.6% X 71.8%). There are no generally accepted standards for measuring particle size distribution from nebulizers and the differences that were observed between methods, which may be related to droplet evaporation, have been described in the literature (55). The results of these experiments are in line with published scintigraphy studies with Aerogen Solo\Ultra in which pulmonary aerosol deposition was 34.1 ± 6.0% of the metered dose (41).

**Supplementary Tables and Figures**

**Supplementary Table 1. Summary of published studies on in vitro inhibition of SARS-CoV-2 by hydroxychloroquine**

| Source | Cell type | Viral input | Type of intervention with hydroxychloroquine | Metric of Viral replication | $IC_{50}$ or viral inhibition* |
|---|---|---|---|---|---|
| Maisonnasse et al.(6) | Vero E6 | MOI 0.01 | Post-infection (1hr) | RT–qPCR in supernatant | 2.2 µM (48 h.p.i), 4.4 µM (72 h.p.i) |
| | Reconstituted Human Airway Epithelium (MucilAir, Epithelix) | MOI 0.1 | Post-infection (1 hr) | RT–qPCR in washing of cells and TCID | No inhibition at 48 h.p.i with highest HCQ concentration that was tested, 10 µM |
| Si et al.(9) | Human Organ-On-A-Chip - microfluidic culture device lined by highly differentiated human primary lung airway epithelium and endothelium | Pseudotyped SARS-CoV-2, MOI not provided | Pre-treatment (24 hr) with drug by perfusing chip's vascular channel followed by infection of chip's airway channel | RT-qPCR in epithelium from chips | No inhibition with highest concentration tested – 1.25 µM (48 h.p.i) |

| | | | | | |
|---|---|---|---|---|---|
| | Huh-7 | Pseudotyped SARS-CoV-2, MOI not provided | Simultaneous infection and treatment | Luciferase activity | ~47% inhibition of viral entry at 1 µM and ~68% inhibition of viral entry at 5 µM (72 h.p.i) |
| Liu et al.(3) | Vero E6 | MOI 0.01 | Pre-treatment (1 hr) | RT–qPCR in supernatant | 4.51 µM (48 h.p.i) |
| | | MOI 0.02 | | | 4.06 µM (48 h.p.i) |
| | | MOI 0.2 | | | 17.31 µM (48 h.p.i) |
| | | MOI 0.8 | | | 12.96 µM (48 h.p.i) |
| Yao et al.(4) | Vero E6 | MOI 0.01 | Post-infection (2 hr) | RT–qPCR in supernatant | 6.14 µM (24 h.p.i) |
| | | | | | 0.72 µM (48 h.p.i) |
| | | | Pre-treatment (2 hr) | RT–qPCR in supernatant | 6.25 µM (26 h.p.i) |
| | | | | | 5.85 µM (50 h.p.i) |
| Touret et al.(5) | Vero E6 | MOI not provided | Pre-treatment (15 min) | RT–qPCR in supernatant | 4.17 µM (48 h.p.i) |
| | Caco-2 | | Pre-treatment (15 min) | RT–qPCR in supernatant | ~68% inhibition at 5 µM and ~58% inhibition at 10 µM |
| Hoffman et al.(2) | Vero E6 | Pseudotyped SARS-CoV- | Pre-treatment (2 hr) | Firefly Luciferase activity as indicator of transduction efficiency | 13.3 µM (18 h.p.i) |
| | Vero E6 +TMPRSS2 | | | | No inhibition with highest concentration |

| | | | | | |
|---|---|---|---|---|---|
| | | 2, MOI not provided | | | tested – 100 µM (18 h.p.i) |
| | Calu-3 | | | | ~47% inhibition at highest concentration tested – 100 µM, $IC_{50}$ reported as 119 µM (18 h.p.i) |
| Rosenke et al.(8) | Vero E6 | MOI 0.01 | Pre-treatment (1 hour) | RT–qPCR in supernatant | 0.16 µM (72 h.p.i) |
| Mulay et al.(7) | Novel 3D alveolar organoid (tested in suspension) and proximal airway air liquid interface culture system produced from human lung tissue obtained from deceased organ donors. | SARS-CoV-2 inoculum of $1x10^4$ $TCID_{50}$ (infection of apical chamber for the proximal airway system) | Pre-treatment (3 hours); treatment of both apical chamber and basement chamber of air liquid interface | RT-qPCR in | 10 µM of HCQ lead to a 2.4-$log_2$ reduction in viral N gene RNA (48 h.p.i), but with variable effects that were donor epithelium-dependent |

* If an $IC_{50}$ value was not provided, other data on viral inhibition that were reported are provided, including data extracted from figures using WebPlotDigitizer.

h.p.i = hours post-infection; HCQ = Hydroxychloroquine; $IC_{50}$ = Half-maximal inhibitory concentration; MOI = Multiplicity of infection; RT-qPCR = reverse transcription quantitative polymerase chain reaction; TCID = Tissue culture infectious dose; $TCID_{50}$ = Median culture infectious dose

**Supplementary Table 2. Dosing and sampling times for pharmacokinetic and tissue distribution profile of HCQ after single PO, IV, and IT administration of HCQS solution in male Sprague Dawley rats**

| Group | Route | n | Human HCQS dose (mg/kg body weight) | Lung HCQS dose (mg/g lung tissue) | Rat HCQS dose* (mg/kg body weight) | Solution Volume (mL/kg) | Solution (mg/mL) | Termination** |
|---|---|---|---|---|---|---|---|---|
| 1a | IV | 3 | 9.9 | | 9.9 | 1.0 | 9.9 | 2 min |
| 1b | IV | 3 | 9.9 | | 9.9 | 1.0 | 9.9 | 24 hr |
| 2a | IT | 3 | - | 0.03 mg/g | 0.18 | 0.2 | 0.9 | 2 min |
| 2b | IT | 2 | - | 0.03 mg/g | 0.18 | 0.2 | 0.9 | 6 hr |
| 2c | IT | 3 | - | 0.03 mg/g | 0.18 | 0.2 | 0.9 | 24 hr |
| 3a | PO | 3 | 13.3 | | 13.5 | 5.0 | 2.7 | 2 min |
| 3b | PO | 3 | 13.3 | | 13.5 | 5.0 | 2.7 | 24 hr |

*Rat weight 0.25 kg and rat lung weight 1.5 g (42). The PO dose was adjusted to 13.5 mg/kg for technical reasons.

**Blood sampling times: Animals terminated at 2 min (pre-dose and termination); animals terminated at 6 hr (pre-dose and termination); animals terminated at 24 hr after IV or IT administration (pre-dose, and post-dose: 2, 10, 20, and 30 min; 1, 2, 4, 6, and 24 hours); animals terminated at 24 hr after PO administration (pre-dose, and post-dose: 5, 15, and 30 min; 1, 2, 4, 6, 8, and 24 hours).

HCQS = Hydroxychloroquine sulfate; IT = Intratracheal; IV = Intravenous; PO = Oral gavage

**Supplementary Table 3. Study Design of Syrian Hamster SARS-CoV-2 Infection Model**

<table>
<tr><th>Group</th><th>N</th><th>IT HCQS*</th><th>IN HCQS*</th><th>SARS-CoV-2 challenge</th><th>Study Assessments</th></tr>
<tr><td>High Dose</td><td>7</td><td>0.072 mg (equivalent to human dose of 120 mg loaded in the nebulizer)</td><td>0.24 mg</td><td rowspan="3">**</td><td rowspan="3">• Weight (daily, days 1-7)<br>• Oral swab for SARS-CoV-2 (day 2, 4, and 7)<br>• Termination (day 7)***</td></tr>
<tr><td>Low Dose</td><td>7</td><td>0.036 mg (equivalent to human dose of 60 mg loaded in the nebulizer)</td><td>0.12 mg</td></tr>
<tr><td>Control</td><td>5</td><td>NA</td><td>NA</td></tr>
</table>

* IT HCQS was administered once daily on days -3, -2, and -1, followed by SARS-CoV-2 challenge on day 0 and IN HCQS 4 hours post-challenge. The HCQS dose was diluted with 0.9% sodium chloride to a total volume of 10 µL for IT dosing and 100 µL for IN dosing.

** Viral challenge consisted of 1:10 dilution of thawed virus stock with phosphate buffered saline (PBS) for a total delivery volume of 50 µL per nostril.

*** At termination on Study Day 7, all animals in all groups were euthanized and the lungs were weighed and fixed in 10% neutral buffered formalin solution. The resulting tissues were forwarded to Histo-Scientific Research Laboratories (Mount Jackson, VA) where they were processed, embedded in paraffin, sectioned and stained with hematoxylin and eosin.

HCQS = Hydroxychloroquine sulfate; IN = Intranasal; IT = Intratracheal

**Supplementary Table 4. Key pharmacokinetic parameters after single PO, IV, and IT administration of HCQS solution in male Sprague Dawley rats**

| Route of administration | Dose of HCQS (mg/kg) | $T_{max}$, range (min) | $C_{max}$ in whole blood (ng/mL) | $C_{max}$ in plasma (ng/mL) | Whole blood $AUC_{0\text{-}6hr}$ (ng*hr/mL) | Whole blood $AUC_{0\text{-}24hr}$ (ng*hr/mL) |
|---|---|---|---|---|---|---|
| IT | 0.18 | 2-10 | 40.7 ± 11.7 | 61.9 ± 35.3 | 44.2 ± 8.1 | 89.3 ± 20.4 |
| IV | 13.3 | 2-10 | 1,664.0 ± 724.6 | 2,230.0 ± 353.4 | 2,115.0 ± 109.2 | 4,323.0 ± 331.5 |
| PO | 9.9 | 240 | 183.3 ± 48.6 | 68.9 ± 22.9 | 955.6 ± 199.4 | 2,320.3 ± 392.5 |

Results are mean ± SD of measurements in 3 animals for each route of administration. PK parameters ($C_{max}$, AUC) are reported with units of ng of hydroxychloroquine base.

AUC = Area under the curve; $C_{max}$ = Maximum concentration; HCQS = Hydroxychloroquine sulfate; IT = Intratracheal; IV = Intravenous; PO = Oral gavage; $T_{max}$ = time of $C_{max}$

**Supplementary Table 5. Comparison of hydroxychloroquine tissue, blood, and plasma concentrations after single PO, IV, and IT administration of HCQS solution in male Sprague Dawley rats**

| Route of administration | Tissue concentration (µg of hydroxychloroquine base / g tissue) | | | | | | | | | | | |
|---|---|---|---|---|---|---|---|---|---|---|---|---|
| | Left heart | | | Right heart | | | Large airways | | | Lung parenchyma | | |
| | 2 min | 6 hr | 24 hr | 2 min | 6 hr | 24 hr | 2 min | 6 hr | 24 hr | 2 min | 6 hr | 24 hr |
| IT (0.18 mg/kg) | 0.72 ± 0.14 | 0.43 ± 0.20 | 0.14 ± 0.05 | 0.61 ± 0.11 | 0.44 ± 0.20 | 0.16 ± 0.08 | 14.61 ± 2.36 | 3.77 ± 0.65 | 1.59 ± 0.70 | 49.53 ± 6.51 | 14.85 ± 0.91 | 3.59 ± 1.95 |
| IV (9.9 mg/kg) | 24.22 ± 0.56 | | 3.17 ± 0.94 | 25.44 ± 5.20 | | 3.18 ± 0.51 | 45.33 ± 5.76 | | 9.42 ± 1.10 | 102.12 ± 17.44 | | 24.11 ± 8.12 |
| PO (13.3 mg/kg) | 1.80 ± 0.43 | | 2.05 ± 0.34 | 1.94 ± 0.39 | | 2.57 ± 0.42 | 3.19 ± 0.71 | | 7.23 ± 0.72 | 9.88 ± 3.43 | | 12.93 ± 3.55 |
| p value (IT vs IV) | <0.001 | | <0.03 | <0.01 | | <0.01 | <0.01 | | <0.001 | <0.02 | | <0.04 |
| p value (IT vs PO) | <0.04 | | <0.01 | <0.02 | | <0.01 | <0.01 | | <0.001 | <0.01 | | <0.02 |

| Route of administration | Concentration (ng/mL) | | | | | |
|---|---|---|---|---|---|---|
| | Whole Blood* | | | Plasma* | | |
| | 2 min | 6 hr | 24 hr | 2 min | 6 hr | 24 hr |
| IT (0.18 mg/kg) | 56.10 ± 10.28 | 5.07 ± 1.82 | 1.72 ± 0.79 | 39.03 ± 4.44 | 9.74 ± 9.6 | 1.89 ± 1.23 |
| IV (9.9 mg/kg) | 2393.33 ± 205.99 | | 73.67 ± 6.99 | 2303.33 ± 417.89 | | 21.93 ± 1.55 |
| PO (13.3 mg/kg) | 71.67 ± 7.51 | | 46.93 ± 8.14 | 22.87 ± 5.69 | | 18.67 ± 1.96 |

* The whole blood and plasma concentrations reported in this table are based on measurements taken at the same timepoint as termination for harvesting tissue (n=3 for each timepoint, except for 6 hr where n=2), and they were used to calculate the tissue:blood ratios of HCQ concentration that are reported in Table 1. Accordingly, the whole blood and plasma concentrations reported in Supplementary Table 5 may appear different that those reported in Figure 2A and Figure 2B, in which the plotted whole blood and

plasma concentrations of HCQ are based on results in 3 rats in each group (see Supplementary Table 2: groups 1b, 2c, 3b) that were terminated only 24 hours post-administration of HCQS solution and thus had whole blood and plasma sampled over 10 different timepoints prior to termination.

IT = Intratracheal; IV = Intravenous; PO = Oral gavage

**Supplementary Table 6. In vitro dose distribution of nebulized HCQS solution from Aerogen Solo\Ultra**

| | HCQS concentration | | | | | |
|---|---|---|---|---|---|---|
| | 20 mg/mL | | 50 mg/mL | | 100 mg/mL | |
| Metered dose loaded in the nebulizer | 1 mL (20 mg) | | 1 mL (50 mg) | | 1 mL (100 mg) | |
| | Dose (mg) | % Recovered | Dose (mg) | % Recovered | Dose (mg) | % Recovered |
| Total dose recovered | 19.2 ± 0.4 | 95.6 ± 2.4 | 49.1 ± 0.4 | 97.8 ± 0.9 | 98.1 ± 1.4 | 98.0 ± 1.5 |
| Aerogen Solo\Ultra (recovered from parts after completion of nebulization) | 9.8 ± 1.2 | 50.8 ± 5.5 | 26.3 ± 3.1 | 53.7 ± 6.2 | 47.2 ± 4.8 | 48.1 ± 5.1 |
| Total HCQS ‘inhaled’ dose (i.e., at the output filter) | 9.1 ± 1.0 | 47.3 ± 5.6 | 21.7 ± 2.8 | 44.2 ± 5.7 | 48.8 ± 5.6 | 49.7 ± 5.3 |
| Total HCQS dose at the exhaust filters | 0.2 ± 0.1 | 1.0 ± 0.9 | 0.5 ± 0.3 | 1.1 ± 1.2 | 1.1 ± 0.3 | 1.1 ± 1.0 |
| Nebulization time | 231 ± 33 sec | | 306 ± 11 sec | | 384 ± 22 sec | |

Data shown are at mean ± SD of 3 nebulizers simulated at each dose with triplicate experiments with each nebulizer ($n$ = 3). Nebulization duration was different ($p<0.001$) with each dose.

HCQS = Hydroxychloroquine sulphate

**Supplementary Table 7. Volumetric droplet diameter distribution of nebulized HCQS solution from Aerogen Solo\Ultra**

| | HCQS concentration | | |
|---|---|---|---|
| | 20 mg/mL | 50 mg/mL | 100 mg/mL |
| Metered dose | 1 mL (20 mg) | 1 mL (50 mg) | 1 mL (100 mg) |
| Laser diffraction experiments | | | |
| Volumetric mean diameter (µm) | 5.0 ± 0.2 | 5.2 ± 0.3 | 4.3 ± 0.3 |
| Percentage of aerosol with volume <5µm | 50.7 ± 2.0 | 47.9 ± 2.8 | 58.6 ± 4.3 |
| Nebulization time | 361 ± 18 | 395 ± 35 sec | 416 ± 9 sec |
| Cascade impaction experiments | | | |
| Emitted dose: total amount of drug assayed from the adaptor to the post-impactor filter (mg) | 13.2 ± 1.2 | 33.8 ± 2.2 | 61.9 ± 5.3 |
| Fine particle fraction of the | 71.8 ± 3.4 | 67.3 ± 3.3 | 71.3 ± 4.7 |

| | | | |
|---|---|---|---|
| emitted dose <5 µM (%) | | | |
| Nebulization time (sec) | 224 ± 26 | 331 ± 17 | 388 ± 30 |

Data shown are at mean ± SD of 3 nebulizers simulated at each dose with triplicate experiments with each nebulizer ($n$ = 3). Nebulization duration was different with each dose ($p<0.005$ for laser diffraction experiments and $p<0.001$ for cascade impaction experiments).

HCQS = Hydroxychloroquine sulfate

### Supplementary Table 8. Summary of Day 1 sensory measures questionnaire results

| Smell | 20 mg (n=2) | 50 mg (n=6) | P (n=2) | Taste | 20 mg (n=2) | 50 mg (n=6) | P (n=2) | Bitterness | 20 mg (n=2) | 50 mg (n=6) | P (n=2) |
|---|---|---|---|---|---|---|---|---|---|---|---|
| No smell | - | 5 | 1 | No taste | - | 1 | 1 | Not bitter | - | 1 | 2 |
| Slight smell, not unpleasant | 1 | 1 | 1 | Slight taste, not unpleasant | - | - | 1 | Slightly bitter, not unpleasant | - | 1 | - |
| Slight smell, unpleasant | 1 | - | - | Slight taste, unpleasant | - | 1 | - | Slightly bitter, unpleasant | - | 3 | - |
| Moderate smell, unpleasant but tolerable | - | - | - | Moderate taste, unpleasant but tolerable | - | 3 | - | Moderately bitter, unpleasant but tolerable | - | 1 | - |
| Strong smell, unpleasant but tolerable | - | - | - | Strong taste, unpleasant but tolerable | 2 | 1 | - | Strongly bitter, unpleasant but tolerable | 2 | - | - |
| Very strong smell, intolerable | - | - | - | Very strong taste, intolerable | - | - | - | Very strongly bitter, intolerable | - | - | - |

20 mg = aHCQS 20 mg dose cohort; 50 mg = aHCQS 50 mg dose cohort; P = Placebo dose cohort

### Supplementary Table 9. Summary of Day 1 acceptability questionnaire results

| **Please rate the taste of the study medication** | | | | **I would be willing to use this study medication on a regular basis** | | | | **The taste of the study medication would not keep me from using it on a regular basis** | | | | **Overall, I would not have a problem using the study medication on a regular basis** | | | | **Please rate how easy it was for you to use the Aerogen device** | | | |
|---|---|---|---|---|---|---|---|---|---|---|---|---|---|---|---|---|---|---|---|
| | **20 mg (n=2)** | **50 mg (n=6)** | **P (n=2)** | | **20 mg (n=2)** | **50 mg (n=6)** | **P (n=2)** | | **20 mg (n=2)** | **50 mg (n=6)** | **P (n=2)** | | **20 mg (n=2)** | **50 mg (n=6)** | **P (n=2)** | | **20 mg (n=2)** | **50 mg (n=6)** | **P (n=2)** |
| Very unpleasant | | - | - | Strongly disagree | - | - | - | Strongly disagree | - | - | - | Strongly disagree | - | - | - | Very difficult | - | - | - |
| Unpleasant | 2 | - | - | Disagree | - | - | - | Disagree | - | - | - | Disagree | - | - | - | Difficult | - | - | - |
| Somewhat unpleasant | - | 5 | - | Somewhat disagree | - | - | - | Somewhat disagree | - | - | - | Somewhat disagree | - | - | - | Somewhat difficult | - | 1 | - |
| Neutral | - | 1 | 2 | Neutral | - | - | 1 | Neutral | - | - | 1 | Neutral | - | - | 1 | Neutral | - | - | - |
| Somewhat pleasant | - | - | - | Somewhat agree | - | 1 | 1 | Somewhat agree | - | - | - | Somewhat agree | - | - | - | Somewhat easy | - | - | - |
| Pleasant | - | - | - | Agree | 1 | 4 | - | Agree | 1 | 3 | 1 | Agree | 1 | 2 | 1 | Easy | 1 | 3 | 1 |
| Very pleasant | - | - | - | Strongly agree | 1 | 1 | - | Strongly agree | 1 | 3 | - | Strongly agree | 1 | 4 | - | Very easy | 1 | 2 | 1 |

20 mg = aHCQS 20 mg dose cohort; 50 mg = aHCQS 50 mg dose cohort; P = Placebo dose cohort

**Supplementary Table 10. Summary of Day 1 acceptability questionnaire results related to exhalation through nose**

| **During the inhalation of the study drug, were you able to exhale through the nose as you were encouraged?** | **20 mg (n=2)** | **50 mg (n=6)** | **P (n=2)** |
|---|---|---|---|
| All of the time | 2 | 3 | 1 |
| Most of the time | - | 3 | - |
| A good bit of the time | - | - | - |
| Some of the time | - | - | 1 |
| A little of the time | - | - | - |
| Hardly any of the time | - | - | - |
| None of the time | - | - | - |

20 mg = aHCQS 20 mg dose cohort; 50 mg = aHCQS 50 mg dose cohort; P = Placebo dose cohort

### Supplementary Table 11. Summary of cough assessment questionnaires

| | Day 1 | | | Day 2 | | | Day 8 | | |
|---|---|---|---|---|---|---|---|---|---|
| | 20 mg (n=2) | 50 mg (n=6) | P (n=2) | 20 mg (n=2) | 50 mg (n=6) | P (n=2) | 20 mg (n=2) | 50 mg (n=6) | P (n=2) |
| **Visual Assessment Scale (0-100) for cough** | **Cough assessment for the time-period between starting the inhalation and until filling out the Day 1 questionnaire (within 3 hours after completion of the inhalation)** | | | **Cough assessment for the time-period between completion of the Day 1 questionnaire and the Day 2 visit** | | | **Cough assessment for the time-period between completion of the Day 1 questionnaire and the Day 8 visit** | | |
| 0 | 1 | 2 | 2 | 2 | 4 | 2 | 2 | 5 | 2 |
| 10 | 1 | 2 | - | - | 2 | - | - | 1 | - |
| 20 | - | 1 | - | - | - | - | - | - | - |
| 30 | - | 1 | - | - | - | - | - | - | - |
| 40-100 | - | - | - | - | - | - | - | - | - |
| | **What was the total duration of cough that you had since starting the inhalation of the study drug?** | | | **What was the total duration of cough that you had since starting the inhalation of the study drug and the Day 2 visit?** | | | **What was the total duration of cough that you had since starting the inhalation of the study drug and the Day 8 visit?** | | |
| All of the time | - | - | - | - | - | - | - | - | - |
| Most of the time | - | - | - | - | - | - | - | - | - |
| A good bit of the time | - | - | - | - | - | - | - | - | - |
| Some of the time | - | 1 | - | - | - | - | - | - | - |
| A little of the time | - | 1 | - | - | 1 | - | - | - | - |
| Hardly any of the time | 1 | 2 | - | - | 1 | - | - | 1 | - |
| None of the time | 1 | 2 | 2 | 2 | 4 | 2 | 2 | 5 | 2 |

20 mg = aHCQS 20 mg dose cohort; 50 mg = aHCQS 50 mg dose cohort; P = Placebo dose cohort

**Supplementary Table 12. Estimation of respiratory tissue HCQ concentration**

| | At 2 minutes post-dose | | | | At 6 hours post-dose | | | | At 24 hours post-dose | | | |
|---|---|---|---|---|---|---|---|---|---|---|---|---|
| Tissue:Blood HCQ concentration in rats after IT administration of HCQS | Large airways | Lung parenchyma | Left heart | Right heart | Large airways | Lung parenchyma | Left heart | Right heart | Large airways | Lung parenchyma | Left heart | Right heart |
| | 265.1 | 900.5 | 12.8 | 10.9 | 771.0 | 3168.6 | 83.5 | 84.5 | 1025.6 | 2473.8 | 93.4 | 107 |
| Observed mean whole blood HCQ concentration in Phase 1 Study after single 50 mg dose of aHCQS | 166.8 ng/mL/0.497 µM | | | | 15.9 ng/mL/0.047 µM | | | | 4.9 ng/mL/0.015 µM | | | |
| Estimated human tissue concentration (µM) | 131.8 | 447.5 | 6.4 | 5.4 | 36.2 | 148.9 | 3.9 | 4.0 | 15.4 | 37.1 | 1.4 | 1.6 |

aHCQS = aerosolized hydroxychloroquine sulfate; HCQ = Hydroxychloroquine, HCQS = Hydroxychloroquine sulfate; IT = Intratracheal

**Supplementary Table 13. Incidence and average severity of microscopic lung findings by treatment group in Golden Syrian hamsters that were treated with IT HCQS on days -3, -2, and -1, followed by SARS-CoV-2 challenge, and IN HCQS 4 hours post-challenge**

| Dose | HCQS High | HCQS Low | Control |
|---|---|---|---|
| Number of Animals Evaluated Microscopically | 7 | 7 | 5 |
| **MICROSCOPIC FINDINGS** | Incidence (Average Severity on a scale of 1-5) | | |
| Accumulation, alveolar macrophages | 7 (1.6) | 7 (1.6) | 5 (1.8) |
| Edema | 7 (1.1) | 5 (1.4) | 4 (1.0) |
| Hemorrhage | 7 (1.4) | 7 (1.4) | 5 (1.2) |
| Hyperplasia, pleural mesothelium | 5 (1.0) | 4 (1.0) | 3 (1.0) |
| Hypertrophy/hyperplasia, bronchiolar epithelium | 7 (2.3) | 7 (2.1) | 5 (2.0) |
| Hypertrophy/hyperplasia, Type II pneumocytes | 7 (3.9) | 7 (3.4) | 5 (3.6) |
| Infiltrate, mononuclear, peribronchiolar/perivascular | 7 (1.9) | 7 (1.7) | 5 (2.0) |
| Inflammation, heterophilic, alveolar | 7 (2.6) | 7 (2.7) | 5 (3.0) |
| Necrosis, bronchiolar epithelium | 7 (1.4) | 7 (1.3) | 5 (1.4) |

| | | | |
|---|---|---|---|
| Syncytia, Pneumocytes | 7 (2.3) | 7 (1.7) | 5 (1.8) |
| Transmigration, vascular, leukocytes | 5 (1.2) | 6 (1.0) | 5 (1.4) |

Average severity grades: 1 = minimal, 2 = mild, 3 = moderate, 4 = marked, 5 = severe.

HCQS = hydroxychloroquine sulfate; IN = intranasal; IT = intratracheal; HCQS High = IT dose 0.072 mg, IN dose 0.24 mg; HCQS Low = IT dose 0.036 mg, IN dose 0.12 mg

**Supplementary Figure 1. Aerogen Solo\Ultra inhalation system and Setup for the dose output breath simulator experiments**

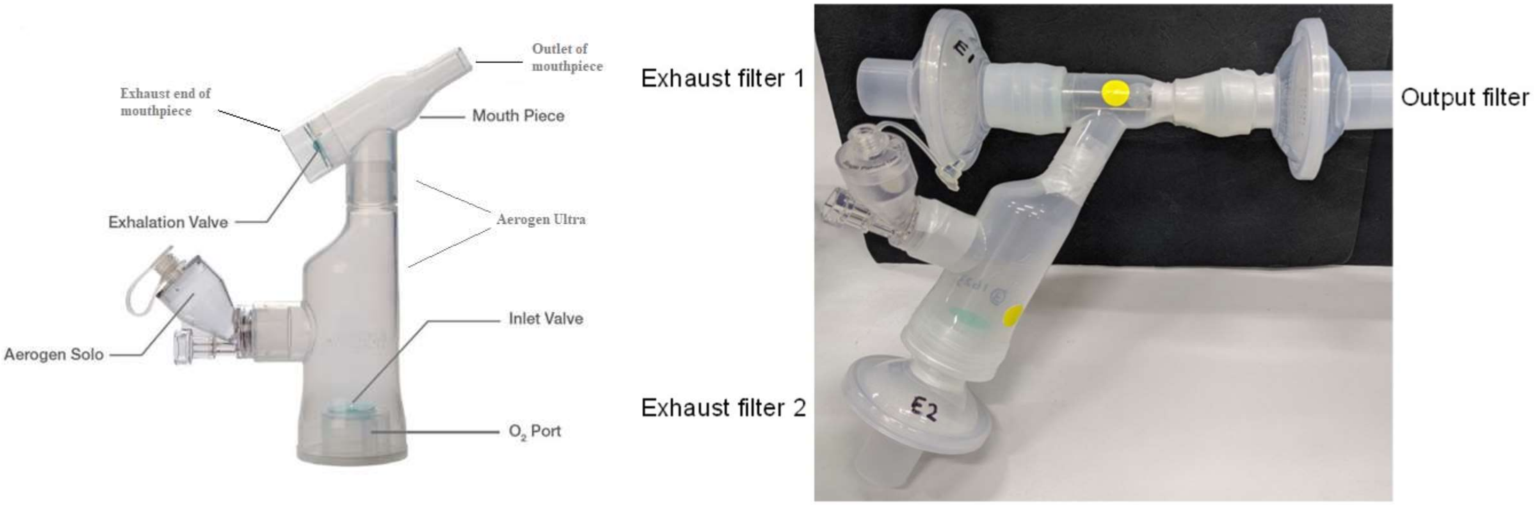

**Supplementary Figure 2. Ratio of HCQ concentration in whole blood vs plasma in the Phase 1 study and rat study**

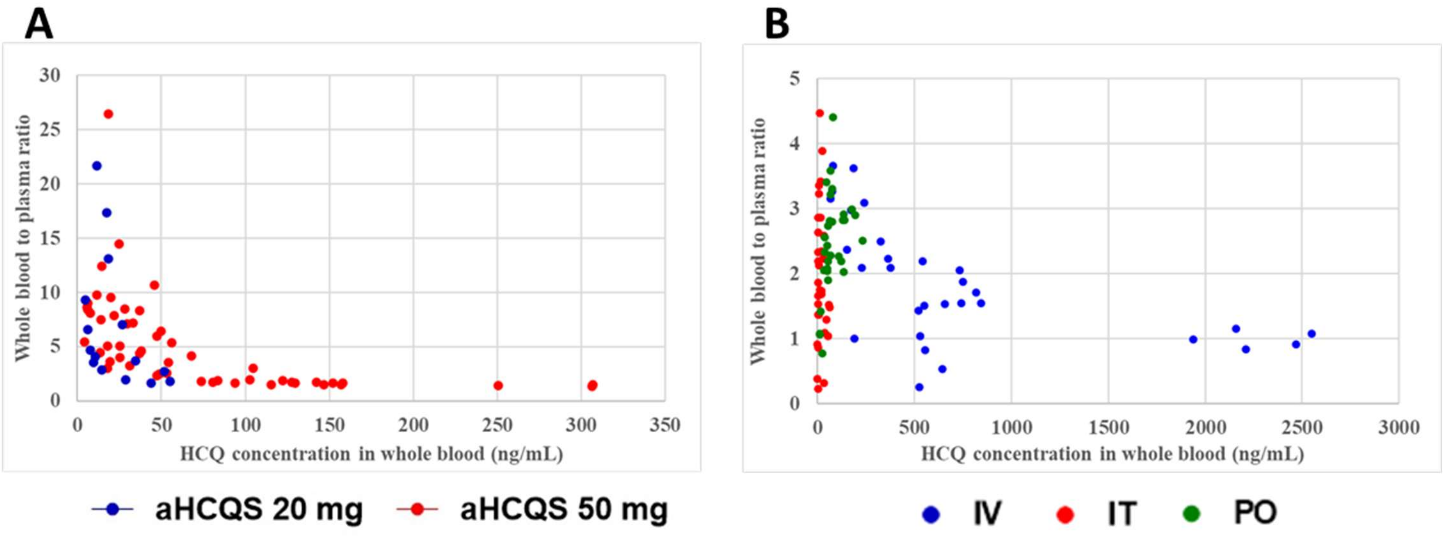

A) Phase 1 study - measurable HCQ concentrations in whole blood and plasma from 8 participants (67 measurements) who received aHCQS. B) Rat study - measurable HCQ concentrations in whole blood and plasma from 20 rats (92 measurements) treated with HCQS (IV 9.9 mg/kg, IT 0.18 mg/kg, PO 13.3 mg/kg).

aHCQS = Aerosolized hydroxychloroquine sulfate; HCQ = Hydroxychloroquine base; HCQS = Hydroxychloroquine sulafte; IT = Intratracheal; IV = Intravenous; PO = Oral

**Supplementary Figure 3. Volumetric droplet diameter distributions of nebulized HCQS solution from Aerogen Solo\Ultra**

**A**

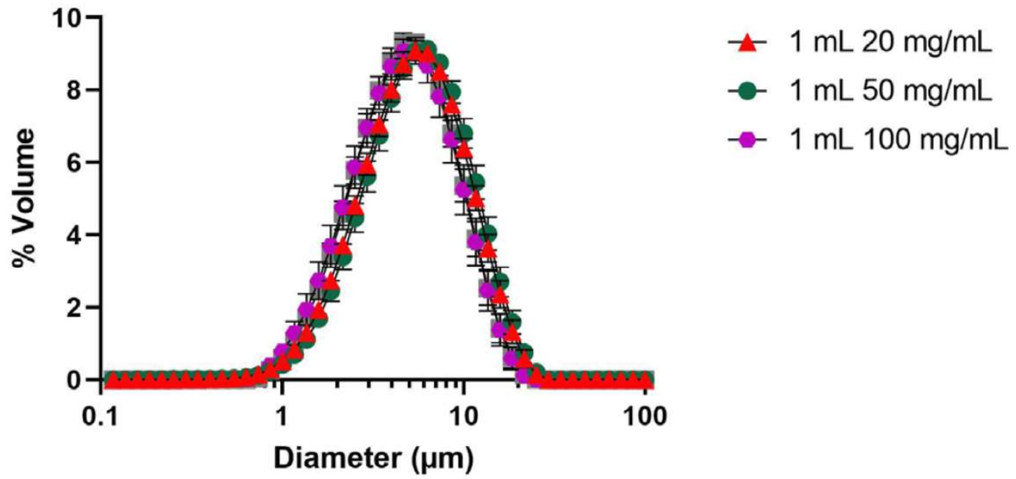

**B**

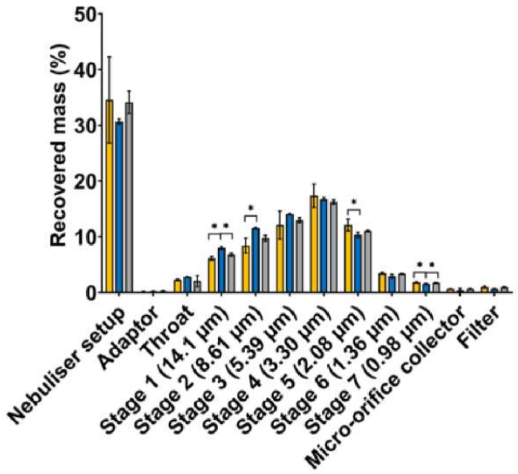

A) Drpolet size distributions measured by laser difraction. Data shown are at mean ± SD of 3 nebulizers simulated at each dose with triplicate experiments with each nebulizer ($n$ = 3). B) Relative doses of HCQS on the various parts of the NGI setup with a metered dose of 1 mL 50 mg/mL HCQS. Data shown are mean ± SD of 3 nebulizers simulated at each dose with triplicate experiments with each nebulizer ($n$ = 3), each nebulizer is marked by a different color.

**Supplementary Figure 4. Pathologic analysis of the lungs of hamsters 7 days post-IN challenge with SARS-CoV-2**

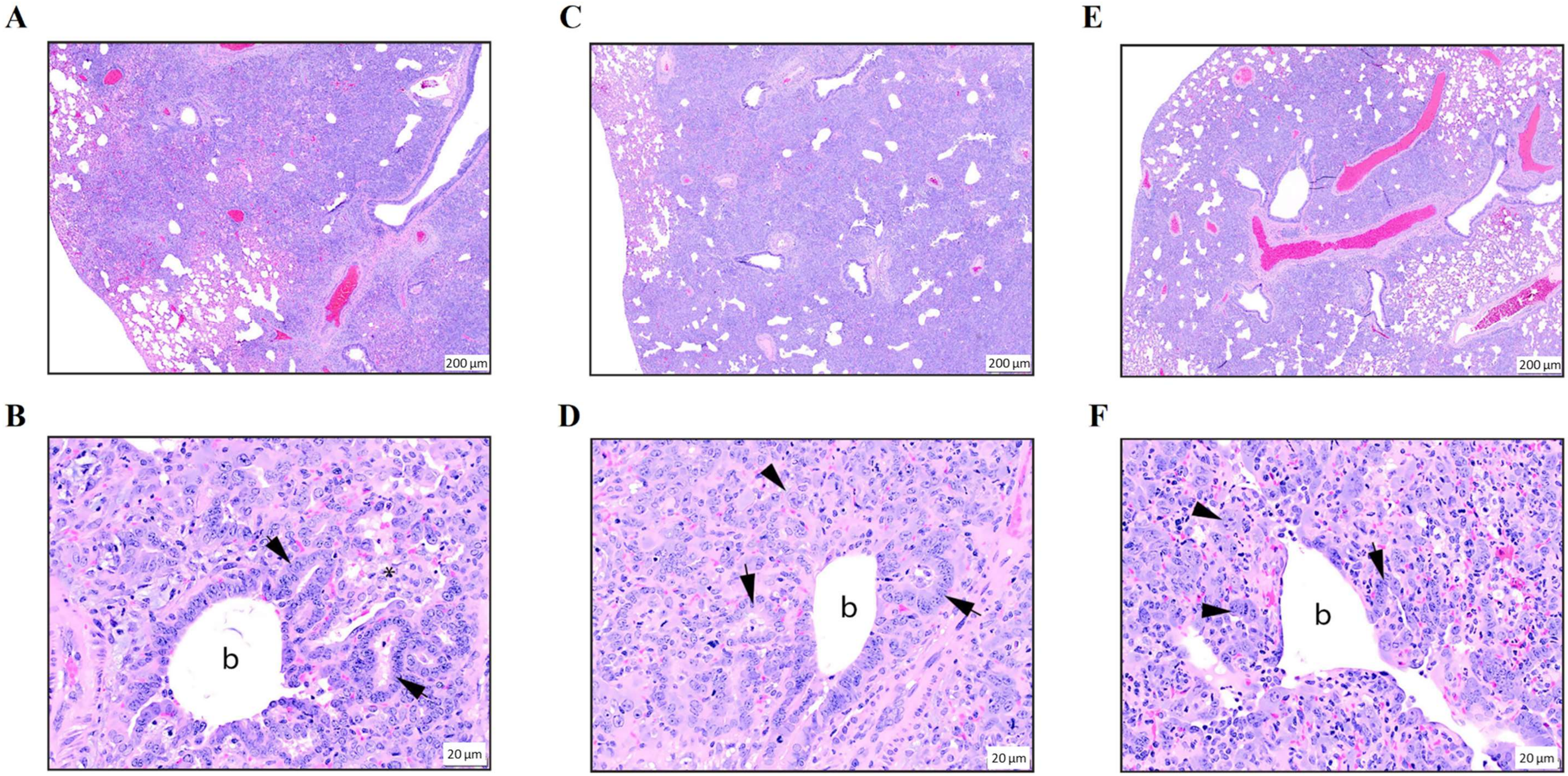

Representative microscopic lung lesions in Syrian hamsters 7 days post-IN challenge with SARS-CoV-2, H&E staining. **High Dose HCQS:** A) Extensive alveolar consolidation in a lung lobe from a hamster treated with high-dose HCQS. Lesions are oriented around bronchi and bronchioles and extend to the pleural surface (20x original magnification). B) Alveoli surrounding a terminal bronchiole (b) are lined by tall, crowded columnar epithelial cells, indicative of bronchiolization (arrows). Foamy alveolar macrophages fill several alveoli (asterisk). Together, these lesions collapse alveolar lumens, which are devoid of air (200x original magnification). **Low Dose HCQS:** C) Extensive alveolar consolidation in a lung lobe from a hamster treated with low-dose HCQS. Lesions are oriented around bronchi and bronchioles and extend to the pleural surface (20x original magnification). D) Alveoli surrounding a terminal bronchiole (b) are lined by tall, crowded columnar epithelial cells, indicative of bronchiolization (arrows). Type II pneumocyte hypertrophy is noted, with occasional syncytial cells (arrowhead). Together, these lesions collapse alveolar lumens, which are devoid of air (200x original magnification). **Untreated control:** E) Extensive alveolar consolidation in a lung lobe from an untreated control hamster. Lesions are oriented around bronchi and bronchioles and extend to the pleural surface (20x original magnification). F) A terminal bronchiole (b) with flattened, attenuated epithelium is surrounded by alveoli lined by tall, crowded columnar epithelial cells, indicative of bronchiolization (arrow). Type II pneumocyte hypertrophy is noted, with occasional syncytial cells (arrowheads). Together, these lesions collapse alveolar lumens, which are devoid of air (200x original magnification).

HCQS = hydroxychloroquine sulfate; IN = intranasal; IT = intratracheal; High Dose HCQS = IT dose 0.072 mg, IN dose 0.24 mg; Low Dose HCQS = IT dose 0.036 mg, IN dose 0.12 mg

49. Fink JB, Ehrmann S, Li J, Dailey P, McKiernan P, Darquenne C, et al. Reducing aerosol-related risk of transmission in the era of COVID-19: An interim guidance

endorsed by the International Society of Aerosols in Medicine. *J Aerosol Med Pulm Drug Deliv.* 2020.

50. American Thoracic Society. Pulmonary function laboratories: Advice regarding COVID-19. https://www.thoracic.org/professionals/clinical-resources/disease-related-resources/pulmonary-function-laboratories.php. Accessed 30 November 2020.

51. Maves RC. CHEST: Management of patients with COVID-19 infection. https://www.chestnet.org/Guidelines-and-Resources/Resources/Management-of-Patients-with-COVID-19-Infection. Accessed 30 November 2020.

52. McGrath JA, O'Sullivan A, Bennett G, O'Toole C, Joyce M, Byrne MA, et al. Investigation of the quantity of exhaled aerosols released into the environment during nebulisation. *Pharmaceutics.* 2019;11(2).

53. U.S. Pharmacopeia. *USP 42 - National Formulary 37, U.S. Pharmacopeial Convention, 2020.* Rockville, MD.

54. Chan JG, Traini D, Chan HK, Young PM, and Kwok PC. Delivery of high solubility polyols by vibrating mesh nebulizer to enhance mucociliary clearance. *J Aerosol Med Pulm Drug Deliv.* 2012;25(5):297-305.

55. Kwong WT, Ho SL, and Coates AL. Comparison of nebulized particle size distribution with Malvern laser diffraction analyzer versus Andersen cascade impactor and low-flow Marple personal cascade impactor. *J Aerosol Med.* 2000;13(4):303-14.
